# A surgery-informed precision approach to determining brain targets for real-time fMRI neurofeedback modulation in chronic pain

**DOI:** 10.1101/2024.05.24.24307873

**Authors:** Dan Liu, Yiqi Mi, Menghan Li, Anna Nigri, Marina Grisoli, Keith M Kendrick, Benjamin Becker, Stefania Ferraro

## Abstract

**Objective:** Despite the promising results of neurofeedback with real-time functional magnetic resonance imaging (rt-fMRI-NF) in the treatment of various psychiatric and neurological disorders, few studies have investigated its effects in acute and chronic pain and with mixed results. The lack of clear neuromodulation targets, rooted in the still poorly understood neurophysiopathology of chronic pain, has probably contributed to these inconsistent findings. In contrast, functional neurosurgery (funcSurg) approaches targeting specific brain regions have been shown to reduce pain in a considerable number of patients with chronic pain, however, their invasiveness limits their use to patients in critical situations. In this work, we sought to redefine, in an unbiased manner, rt-fMRI-NF future targets informed by the long tradition of funcSurg approaches.

**Methods:** using independent systematic reviews, we identified the targets of the rt-fMRI-NF (in acute and chronic pain) and funcSurg (in chronic pain) studies and characterized their underlying functional networks using a subset of high spatial resolution resting-state fMRI data (7T MRI data from the Human Connectome Project). After applying principal component analysis to reduce the number of identified networks, we performed a quantitative functional and anatomical annotation of these networks with a large-scale meta-analytic approach. Finally, we characterized the functional networks, defining their degree of overlap with canonical intrinsic brain networks (default mode, salience, and somatosensory) and their neurotransmitter profile.

**Results:** As expected, the rt-fMRI-NF and funcSurg targets were different, except for the middle cingulate cortex, and showed different characteristics in terms of their functional connectivity. Our findings indicate that targets of rt-fMRI-NF primarily encompass hubs within the default mode network and, to a lesser extent, within the salience network. In contrast, funcSurg targets predominantly involve hubs within the sensorimotor system (primarily the motor system), with less robust involvement of the salience network. Notably, 3 out of 4 derived funcSurg rs-fMRI networks correlated significantly with the distribution map of noradrenaline transporters, further supporting the functional relevance of the funcSurg networks as targets for the treatment of chronic pain.

**Conclusion:** Key hubs of the sensorimotor networks, in particular the motor system, may represent promising targets for the therapeutic application of rt-fMRI-NF in chronic pain in particular in neuropathic pain patients. Our results also suggest that the antinociceptive effects of the funcSurg approaches could be, at least partially, linked to the restoration of abnormal noradrenergic system activation.

## 1. Introduction

Chronic pain, defined as pain that persists or recurs for more than three months (Raja et al. 2020), imposes a very high burden on individuals, families, and society. With prevalence estimates ranging from 18% to 32% of the population in various regions, including China (Yongjun et al. 2020), the United States (Yong, Mullins, and Bhattacharyya 2022), Europe (Breivik et al. 2006), and Africa (Kamerman et al. 2020), the social and economic costs of chronic pain are enormous worldwide exceeding the cost of cancer, diabetes, and heart disease combined (Dydyk and Conermann 2024). Indeed, not only does chronic pain incur significant direct long-term costs related to health care and medical management, but it also results in considerable indirect costs from individuals’ reduced ability to participate fully in social roles, impacting work productivity and family dynamics (Landefeld et al. 2017) with apparent multi-level cascading effects.

Chronic pain is now considered to be a disorder of the central nervous system. However, the central nervous system should not be considered the only key player in the initiation and/or maintenance of chronic pain, as previously argued (Sullivan et al. 2013). The variety of chronic pain conditions and the different degrees of associations with comorbidities with emotional disorders (e.g., depression and anxiety) in around 35% of cases (Asmundson and Katz 2009; Cheung et al. 2020), suggest that chronic pain is not a single nosological entity but possibly a constellation of brain disorders with shared abnormalities. To date, countless clinical and preclinical studies have demonstrated dysregulations in at least three broad functional networks: the mesocorticolimbic system, involved in reward and punishment and emotions and pain processing (Serafini, Pryce, and Zachariou 2020), the salience network (Kim et al. 2018), orchestrating brain activity in response to changes in the external and internal environment with a prominent role in autonomic processes (Ferraro et al. 2022b) and the default mode network (DMN), involved in self-referential activity and the modulation of pain and emotions (Baliki et al. 2008; Yu et al. 2014). Although a solid brain-based model of chronic pain has not yet been established, this knowledge is revolutionizing the treatment of chronic pain in particular with studies that seek to modulate highly specific brain regions in an attempt to control pain levels.

Functional neurosurgery (funcSurg), which includes techniques such as deep brain stimulation (DBS) and ablation of specific brain areas with different approaches (with surgical treatment, but also with the newer gamma-knife and MRgFUS or magnetic resonance-guided focused ultrasound), has a long tradition in treating patients, particularly with severe cases of chronic pain. Remarkable studies have provided evidence that stimulation or lesion of selected targets such as specific nuclei of the thalamus [notably the mediodorsal, ventral posterior lateral and medial (VPL/VPM), central lateral (CL) nuclei], or of the midbrain [periaqueductal gray (PAG)/periventricular gray (PVG) matter] or regions of the middle cingulate cortex (MCC), or the primary motor cortex (Nguyen et al. 2011) can reduce pain levels and alleviate the emotional suffering associated with pain in a good proportion of patients. More specifically, both thalamic/midbrain and MCC DBS approaches have been shown to have a long-term success rate in around 40% of the patients (Bittar et al. 2005; Boccard et al. 2017; Cruccu et al. 2007) with a recent meta-analysis (Frizon et al. 2020) underlying different efficacy of thalamic DBS depending on the targets and the type of chronic pain. However, despite the remarkable results, the invasiveness of these brain-based interventions has inherently prevented their translation into routine clinical practice in the treatment of chronic pain, limiting their use to very severe cases.

In recent years, neurofeedback as a non-invasive form of brain modulation with a highly promising therapeutic potential has gained increasing interest. The recent development of real-time functional magnetic resonance imaging feedback (rt-fMRI-NF), leveraging on the relatively high spatial resolution of fMRI, can provide feedback on the activity of precise brain regions or networks (Grech et al. 2008). Training-induced changes in regional activity and pathways have been shown to induce specific changes in cognitive and emotional processes associated with these neural systems (Yao et al. 2016; Zhao et al. 2019), including acute pain (Emmert et al. 2014; Rance et al. 2014a,b). Notably, rt-fMRI-NF has shown promising results in the treatment of several psychiatric and neurological disorders, such as, among others, depression (Hamilton et al. 2016), post-traumatic stress disorder (Chiba et al. 2019; Zweerings et al. 2020), obsessive-compulsive disorder (Rance et al. 2023; Scheinost et al. 2013), addiction (Hanlon et al. 2013), and Parkinson’s disease (Subramanian et al. 2011, 2016). These findings provide a solid theoretical and technical foundation for the development of rt-fMRI-NF in treating chronic pain conditions. So far, only two studies have investigated the effects of rt-fMRI-NF in chronic pain patients (DeCharms et al. 2005; Guan et al. 2015), and a few more have examined its impact on induced pain in healthy subjects (Emmert et al. 2014; Rance et al. 2014a,b). While all these studies have provided evidence for the ability of individuals to learn to modulate the activity of specific brain regions with this technique, at the behavioral level, they have provided mixed results. Specifically in chronic pain, the first seminal study (DeCharms et al. 2005) showed that patients learned to regulate the activity of the MCC (reported as anterior cingulate cortex in the article) and reduced the level of chronic pain. However, this study was limited by including a small number of patients and the absence of a robust control condition. Moreover, these results were not replicated in a subsequent study of a larger sample of patients (Guan et al. 2015). Interestingly, a more recent study (Guan et al. 2015), employing a double-blind, randomized study, showed that 6 out of 8 postherpetic neuralgia patients were able to learn to modulate the activity of the anterior cingulate cortex, leading to a decrease in the perception of pain.

It is very plausible that the failure to replicate the results obtained in the study of deCharms et al. (2005) (Guan et al. 2015), and the lack of clear behavioral effects in acute pain (DeCharms et al. 2005; Emmert et al. 2014; Rance et al. 2014a,b), have induced a skepticism toward the potential role of rt-fMRI-NF in chronic pain management thus limiting studies in this direction. However, the few findings suggest that it may be premature to dismiss rt-fMRI-NF in chronic pain. Rigorously controlled studies, with larger sample sizes and brain targets defined in an unbiased manner within the many areas implicated in chronic pain (Tan and Kuner 2021), are urgently needed before concluding that this noninvasive, side-effect-free technique is ineffective in the treatment of chronic pain. In this scenario, it is imperative to provide an unbiased definition of the brain areas or networks to be targeted with rt-fMRI-NF in pain conditions. Until now, rt-fMRI-NF targets have been chosen based on the activity induced during acute pain; however, it is well-known that chronic pain is different from acute pain (Martucci, Ng, and Mackey 2014), being characterized by major neuroplastic reorganizations (Kuner and Kuner 2020). Thus, a reassessment of the areas and underlying circuits that might be at the basis of the antinociceptive effects in patients with chronic pain is necessary. In this context, it is crucial to explore innovative approaches that are informed by therapeutic methods.

Building on this imperative for innovation, one promising direction is the use of rt-fMRI-NF informed by the targets employed by functional surgery (funcSurg) approaches for chronic pain management. Against this background, we capitalized on funcSurg studies to identify critical hubs and networks as possible targets for the application of rt-fMRI-NF in the treatment of chronic pain. Through careful systematic reviews, we first characterized the targets of rt-fMRI-NF (in acute and chronic pain conditions) and funcSurg (in chronic pain conditions) approaches. Next, we identified the functional networks underlying these targets by investigating a subset (30 healthy participants) of the 7T resting-state fMRI (rs-fMRI) dataset of the Human Connectome Project (HCP-Young Adult) (Smith et al. 2013; Van Essen et al. 2013). As a final step, we characterized the obtained functional networks to define their membership of canonical rs-fMRI networks, and their meta-analytic [employing “decoder” function in Neurosynth (Yarkoni et al. 2011)], and neurotransmitter [employing neurotransmitter receptor maps in Juspace (Dukart et al. 2020)] profiles. With this approach, we were able to identify the functional networks underlying the targets used in rt-fMRI-NF studies and the antinociceptive effects of funSurg approaches, allowing us to unbiasedly revise the brain targets for use in future rt-fMRI-NF studies.

## 2. Methods

### 2.1. Records selection

#### 2.1.1. rt-fMRI-NF records selection

The final literature search was conducted in August 2023 (starting from January 2000) and undertaken by one reviewer (LD) using the following strings: [(‘neurofeedback’ OR ‘realtime’) AND (‘fMRI’ OR ‘functional magnetic resonance imaging’ OR ‘functional MRI’) AND ‘pain’] in Web of Science, PubMed, Scopus. Records were screened by two independent reviewers (LD and YM) based on the following criteria: (1) written in English and peer-reviewed; (2) reporting an original study (excluding reviews or commentaries and not duplicated results); (3) using only rt-fMRI-NF training paradigm to modulate the level of pain in healthy participants or chronic pain patients. For the included records, the first reviewer (LD) extracted: (1) demographic information (i.e., population, sample size, and age and gender distribution); (2) feedback characteristics (method of localization of the target and type of feedback); (3) suggested conditioning strategy and direction of request for modulation of target activity (increased or decreased); (4) experimental design (i.e., number of training sessions, use of a control group or control condition); (5) coordinates of the brain targets in the standardized space [Talairach & Tournoux (TAL) (Talairach 1988) or Montreal Neurological Institute (MNI) (Evans et al. 1993)]; (6) behavioral effects on pain level. The second reviewer (YM) independently validated the extracted data: when discrepancies were observed, they were jointly discussed with the senior author of the paper (SF) to reach a final consensus.

Considering that several nomenclatures and subdivisions exist for brain areas, particularly for the cingulate cortex (Vogt 2009), we decided to identify the target regions uniquely, thus independently from the nomenclature given by the Authors, relying on the Automated Anatomical Labelling Atlas 3 (AAL3) (Rolls et al. 2020) and the probabilistic atlas of the insula cortex (Faillenot et al. 2017). Moreover, the original coordinates expressed in the TAL space were converted to MNI stereotactic space using GingerALE (3.0.2).

#### 2.1.2. Functional surgery records selection

To identify the targets of funcSurg approaches in chronic pain, we relied on the most recent meta-analyses in the field by selecting articles from the cited studies. For this purpose, we searched for meta-analyses published from January 2000 to August 2023 in Web of Science, PubMed, and Scopus with the following keywords: [(DBS OR deep brain stimulation OR cingulotomy OR thalamotomy) AND (pain OR chronic pain OR tumor pain) AND (meta-analysis OR review)]. In our work, we did not consider studies employing motor cortex stimulation to treat pain because the use of electrodes covering a vast portion of the motor cortex prevented the identification of relatively precise localization (Lavrov et al. 2021). In addition, because rt-fMRI-NF applies only to gray matter areas, we excluded works that targeted exclusively white matter (such as the posterior limb of the internal capsule) (Nguyen et al. 2011).

Next, to identify recently published articles, we performed an additional literature search in the same databases from the publication date of the identified meta-analysis until August 2023 employing the following keywords: [(DBS OR deep brain stimulation) AND (pain OR chronic pain OR tumor pain)] for DBS; [(cingulotomy) AND (pain OR chronic pain OR tumor pain)] for cingulotomy; [(thalamotomy OR MRgFUS OR Gamma Knife) AND (pain OR chronic pain OR tumor pain)] for thalamotomy. In cases where recent meta-analyses were not available, such as in the case of thalamotomy, we performed the relevant literature search from January 2000 until August 2023. Among the identified papers (from the already published meta-analyses and the more recent studies), records were then selected based on the following criteria: published after 2000, treating at least 8 patients, with a clear description of the target landmarks, resulting in pain relief in at least 40% of patients within the longest follow-up.

Since our interest was in defining the location of the target, where authors of a study localized the target based on a previous study, we utilized the study that reported more detailed information about its location. In addition, when the technique was developed and refined in the same region by the same group, assuming that the latest development was a better target than the previous one, as in the case of thalamotomy with MRgFUS (Gallay et al. 2023), we used the description of the last published work. Moreover, if a paper reported multiple targets in identical regions, we computed their center of mass (Gallay et al. 2023; Strauss et al. 2018).

For each included record, the first reviewer (LD) extracted (1) demographic information, (2) type of chronic pain, (3) behavioral effects on pain level, and (4) information relevant to identifying the coordinates of the target. The second reviewer (YM) independently validated the extracted data. When discrepancies were observed, they were jointly discussed with the senior author of the paper (SF) to reach a final consensus. Also in this case, we decided to identify the cortical target regions uniquely, thus independently from the nomenclature given by the Authors, relying on the Automated Anatomical Labelling Atlas 3 (AAL3) (Rolls et al. 2020). Based on the collected information relative to the localization of the target, three senior researchers (SF, AN, and LM in the acknowledgment note) computed independently the coordinates of each target in the MNI space with MANGO v4.1 (http://rii.uthscsa.edu/mango/) employing the MNI template ICBM 2009b Nonlinear Symmetric template [spatial resolution: 0.5x0.5x0.5mm; mni icbm152 t1 tal nlin sym 09b hires. nii, ICBM 152 Nonlinear atlases (2009) – NIST (mcgill.ca)]. Then, for each target, after verifying that the distance between the points located by the first reviewer relative to the second and third reviewers was not greater than the arbitrary distance of 5 mm, the center of mass of the located points was calculated. As the last step, due to the complexity of the thalamic anatomy and the inherent difficulties in defining the coordinates based on the text of the selected studies, we verified the localization of the extracted coordinates on the in-vivo high-resolution structural-MRI human thalamic atlas (Saranathan et al. 2021). If the reported coordinates were not consistent with the location of the thalamic nuclei of interest, we first wrote to the Authors of the paper to ask for clarifications and, in case of no responses within one month, we used the coordinates of the centroid of the putative thalamic nucleus computed on the same atlas.

### 2.2. Identification of the rs-fMRI networks underlying the target regions

To identify the underlying rs-fMRI networks, Marsbar (v 0.44) was used to create 6 mm (for cortical targets) and 3 mm (for subcortical targets) radius regions of interest (ROIs) centered in the identified MNI coordinates. The 3 mm radius for the subcortical ROIs was chosen because it avoided the overlap between the ROIs from different but nearby regions.

To clarify, the targets for rt-fMRI-NF were always on either the left or right side of specific structures, while the targets for funcSurg were always bilateral. Therefore, when computing the functional connectivity for rt-fMRI-NF ROIs, we employed the single lateralized ROI as a seed. In contrast, for funcSurg ROIs, we employed the left and right ROIs together as seed. Using CONN toolbox v22a (www.nitrc.org/projects/conn) (Nieto-Castanon 2020), we calculated the seed-based functional connectivity of each ROI (as seed for rt-fMRI-NF) or a couple of ROIs (for funcSurg) employing rs-fMRI data from 30 subjects (age: M = 29.17 ys, SD = 3.32 ys; 20 females) of the Young Adult HCP dataset (Smith et al. 2013; Van Essen et al. 2013; for details, see https://www.humanconnectome.org/hcp-protocols-ya-7t-imaging). As declared by HCP, all participants provided written informed consent to the study and the sharing of de-identified data. For each subject, we employed the 4 rs-fMRI data runs acquired at 7T (900 volumes per run, 1.6 mm isotropic voxels, TR = 1000 ms, TE = 22.2 ms, flip angle = 45 degrees, FOV = 208 × 208 mm) (Smith et al. 2013; Uğurbil et al. 2013) and which have been already preprocessed (HCP filename: ‘rfMRI hp2000 clean.nii.gz’). The HCP preprocessing steps comprised gradient nonlinearity-induced distortion correction, rigid body head motion correction, EPI image distortion correction, co-registration between the fMRI and structural data, normalization to MNI space, high-pass filtering, brain masking (Glasser et al. 2013), and independent component analysis based artifact removal of noise components (Salimi-Khorshidi et al. 2014). As structural MRI data, we used 3T T1-weighted images re-sampled at 1.6 mm resolution provided by the HCP for the use with 7T rs-fMRI dataset (filename: ‘T1w restore.1.6.nii.gz’).

Using CONN toolbox v21a (www.nitrc.org/projects/conn), the preprocessed fMRI data from HCP underwent the following preprocessing and denoising steps: segmentation, artifact identification using Artifact Detection Tools (ART), smoothing (FWHM = 6 mm), and standard denoising pipeline (aCompCor). For each ROI or a couple of ROIs, seed-based connectivity (SBC) maps were estimated as Fisher-transformed bivariate correlation coefficients (weighted-GLM) (Nieto-Castanon 2020). Multivariate parametric statistics with random effects across subjects and sample covariance estimation across multiple measurements were applied (cluster threshold: p<0.05 cluster-level, p-FDR corrected; voxel threshold: p<0.001 uncorrected).

### 2.3. Principal components analysis of the identified rs-fMRI networks (PCA rs-fMRI-derived maps)

Separately for the rt-fMRI-NF and the funcSurg targets, we performed a dimensionality reduction with Principal Components Analysis (PCA) of the obtained rs-fMRI maps (8 for rt-fMRI-NF and 8 for funcSurg). This step was crucial for identifying the most significant patterns within the functional connectivity networks. To achieve this, we utilized the PCA class from the *scikit-learn* library in Python (version 3.11.7). For both analyses, we retained only the first components that explain a cumulative variance of at least 80%. Then, we built maps that reflect the information captured by these principal components. These new maps are referred to as PCA rs-fMRI-derived maps.

Next, we asked whether the PCA rs-fMRI-derived maps significantly overlapped with the salience, sensorimotor, and default mode (DMN) networks (herein after named canonical rs-fMRI networks). To this aim, we used a null map approach (Burt et al. 2020) as described in Ferraro et al. (2022), employing the rs-fMRI network maps available at http://findlab.stanford.edu/research.html (Shirer et al. 2012). Briefly, we generated 1000 surrogate maps for each canonical rs-fMRI map (i.e., salience, DMN, and sensorimotor). Then, we computed the null distribution of the overlap scores, computing the number of overlapping voxels between the PCA rs-fMRI-derived map of interest and each of the surrogate maps. Using this null distribution, we then obtained a p-value testing the hypothesis that the overlap between the PCA rs-fMRI-derived map and the canonical rs-fMRI map was greater than expected by chance. To preserve spatial autocorrelation (Markello et al. 2021), the surrogate maps were generated employing the algorithm implemented in Neuromaps (Markello et al. 2022).

### 2.4. Neurosynth decoding of PCA rs-fMRI-derived maps

To identify the cognitive and anatomical terms associated with the PCA rs-fMRI-derived maps, we performed the functional decoding employing the “decoder” function in Neurosynth (https://neurosynth.org/) (Yarkoni et al. 2011). This function decodes cognitive states and anatomical areas from brain images by analyzing the spatial correlation between the map of interest and meta-analytic activation maps in its database. For each PCA rs-fMRI-derived map, the Neurosynth decoder produced a ranked list of terms and their corresponding correlation coefficients. For interpretation purposes, we retained the top 20 terms.

### 2.5. Neurosynth meta-analytic brain maps of pain and chronic pain terms

To have an unbiased representation of the brain activity during pain and chronic pain processing, we employed Neurosynth (https://neurosynth.org/) (Yarkoni et al. 2011) to obtain meta-analytic maps of the terms ‘pain’ and ‘chronic pain.’ More specifically, for each term, we obtained the uniformity test map, which represents the uniform distribution of brain activity associated with that term, thus showing the extent to which each voxel is consistently activated in studies employing the specific word ‘pain’ or ‘chronic pain’. Next, as already done for PCA rs-fMRI-derived maps, we ought to determine which canonical rs-fMRI networks the meta-analytic brain maps belong to. Also, in this case, we determined if the identified meta-analytic maps significantly overlapped with the selected canonical rs-fMRI maps (i.e., salience, DMN, and sensorimotor) employing a null map approach (Burt et al. 2020), as described in the previous section.

### 2.6. Neurotransmitter receptors profiling of PCA maps

To estimate the spatial distribution of neurotransmitter receptors within each PCA rs-fMRI-derived map, we employed the JuSpace toolbox version 1.5 (available at https://github.com/juryxy/JuSpace; (Dukart et al. 2020)). This toolbox allowed the estimation of spatial correlations between our PCA rs-fMRI-derived maps and neurotransmitter receptor distribution maps obtained through nuclear imaging techniques These neurotransmitter receptors’ linearly rescaled maps are derived from different studies that report the average receptor maps observed in groups of healthy participants. Overall, the following neurotransmitter and neuromodulatory systems were investigated: gamma-aminobutyric acid type A (GABA) (Dukart et al. 2018) serotonin 5-hydroxytryptamine receptor subtype 1a (5-HT1a) (Hansen et al. 2022; Savli et al. 2012) and 1b (Beliveau et al. 2017; Savli et al. 2012) serotonin transporter (SERT) (Beliveau et al. 2017; Savli et al. 2012), dopamine D2 (D2) (Alakurtti et al. 2015), dopamine transporter (DAT) (Dukart et al. 2018), noradrenaline transporter (NAT) (Hesse et al. 2017), and opioid (MU and KappaOp) (Hansen et al. 2022; Kantonen et al. 2020; Shokri-Kojori et al. 2022).

Using JuSpace, the PCA rs-fMRI-derived maps, and the neurotransmitter receptor maps were parcellated into 116 regions according to the Automated Anatomical Labeling (AAL) atlas, from which mean regional values were extracted. Adjusting for spatial autocorrelation through partial correlation with grey matter probability estimates, Spearman’s correlation (Fisher’s z-transformed value) was computed between each PCA rs-fMRI-derived map and each neurotransmitter receptor map. Then, the exact p-value of each correlation was calculated by generating randomly permuted neurotransmitter receptor maps (1000) that maintain the same spatial autocorrelation as the original map. These permuted maps were then used to create a null distribution, against which each PCA rs-fMRI-derived map is compared to determine if the observed correlation was significant (p-value<0.0017, Bonferroni correction for 29 statistical tests to control for multiple comparisons).

## 3. Results

### 3.1. Records selection

#### 3.1.1. rt-fMRI-NF records selection

The literature search (see Figure 1) resulted in 202 articles after removing the duplicates. Based on the title and the abstract, 192 articles were excluded, leaving 10 papers. After the entire reading, 5 papers were excluded. The remaining 5 records investigated a total of 84 healthy individuals (DeCharms et al. 2005; Emmert et al. 2014; Guan et al. 2015; Rance et al. 2014a,b) and 28 chronic pain patients (DeCharms et al. 2005; Guan et al. 2015). These studies have employed rt-fMRI-NF to evaluate the feasibility and the behavioral effects of modulating specific brain targets on laboratory-induced acute pain (DeCharms et al. 2005; Emmert et al. 2014; Guan et al. 2015; Rance et al. 2014a,b), and chronic pain perception (DeCharms et al. 2005).

**Figure 1:**
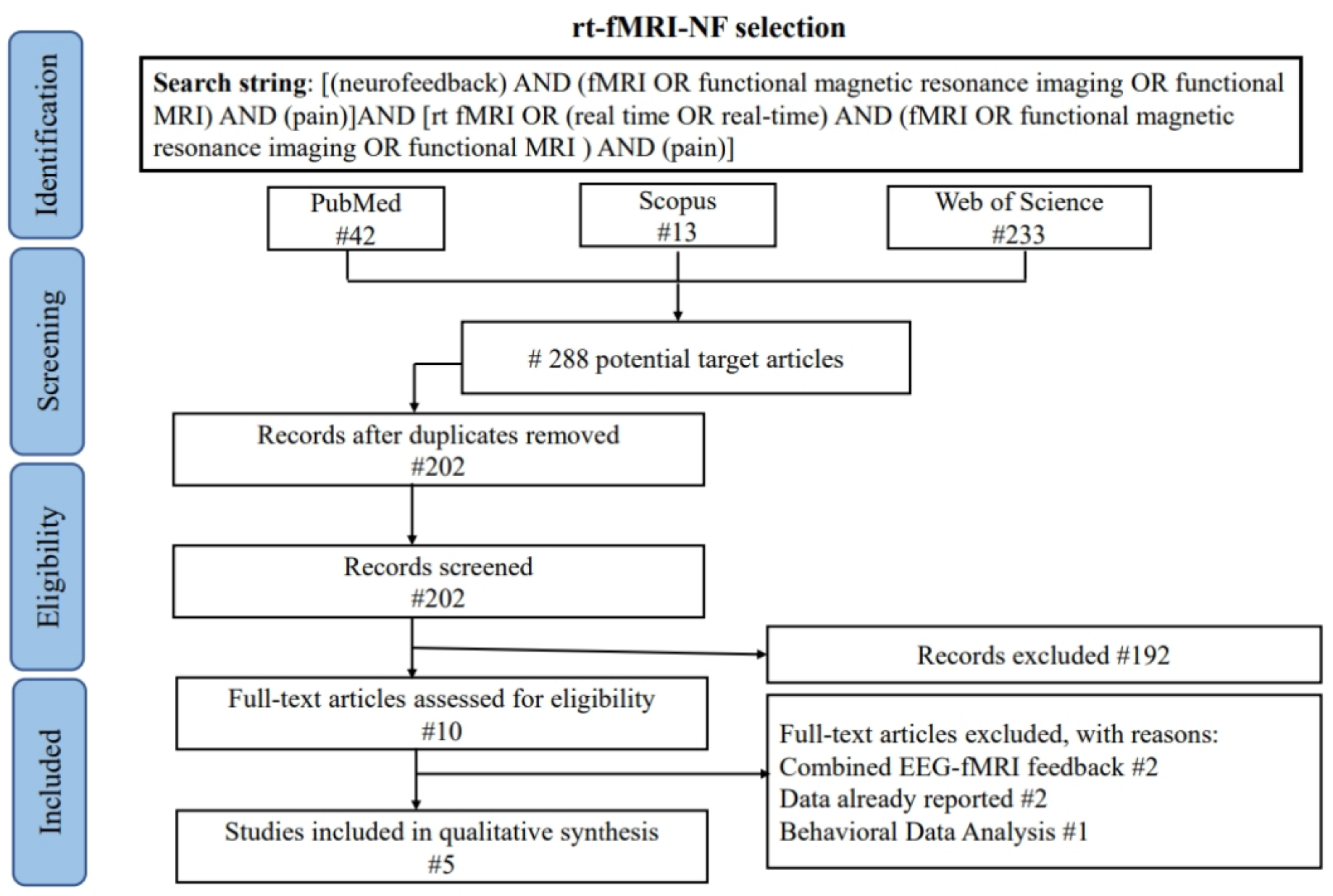
Flowchart of the screening process of the rt-fMRI-NF studies according to the PRISMA guideline.

The MNI coordinates of the targets (original coordinates expressed in the TAL stereotactic space in DeCharms et al. 2005; Guan et al. 2015; Rance et al. 2014a,b) were identified in the MCC (DeCharms et al. 2005), in the supracallosal (Emmert et al. 2014), subgenual (Rance et al. 2014a,b) and pregenual (Guan et al. 2015) anterior cingulate cortex (ACC), in the posterior insula cortex (pIns) (posterior longitudinal gyrus) (Rance et al. 2014a,b), and in the anterior insula cortex (aIns) (middle short gyrus) (Emmert et al. 2014). Despite all the studies reporting that the participants were able to learn to modulate the activity of the target, only 2 also demonstrated significant behavioral effects. These two studies, which investigated the modulation of acute pain in healthy participants and chronic pain in chronic pain patients, provided evidence of the potential of rt-fMRI-NF in pain management (DeCharms et al. 2005; Guan et al. 2015). Specific information on the 5 included studies is shown in Table 1 and identified coordinates in Table 2.

**Table 1:**
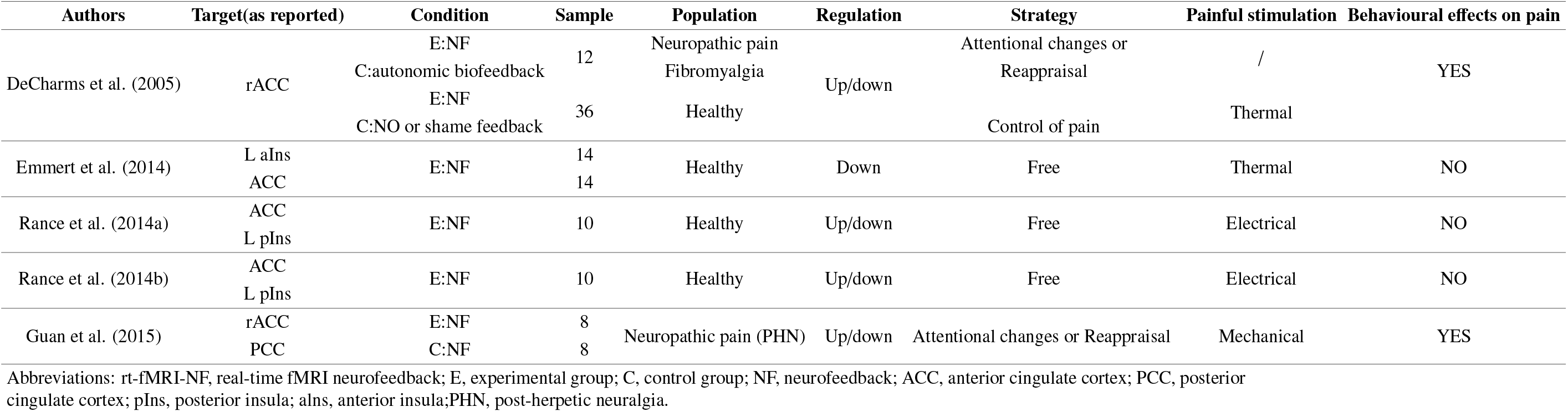
Selected rt-fMRI-NF studies.

**Table 2:** Coordinates of the targets of the rt-fMRI-NF studies.

| Author | Target<br>(as reported) | Target area<br>(AAL3) | Radius<br>(mm) | MNI coordinates |  |  |
| --- | --- | --- | --- | --- | --- | --- |
| DeCharms et al. (2005) | R rostral ACC | R MCC | 6 | 4 | 22 | 30 |
| Emmert et al. (2014) | L aIns | L aIns | 6 | -38 | 4 | 3 |
|  | L ACC | L ACCsup | 6 | -1 | 21 | 26 |
| Rance et al. (2014a) | rostral ACC | L ACCsub | 6 | 0 | 37 | -2 |
|  | L pIns | L pIns | 6 | -42 | -19 | 11 |
| Rance et al. (2014b) | L rostral ACC | L ACCsub | 6 | -3 | 35 | -1 |
|  | L pIns | L pIns | 6 | -45 | -4 | -7 |
| Guan et al. (2015) | R rostral ACC | R ACCpre | 6 | 1 | 41 | 1 |

Abbreviations: rt-fMRI-NF, real-time fMRI neurofeedback; ACC, anterior cingulate cortex; MCC, middle cingulate cortex; ACCsub, anterior cingulate cortex, subgenual; ACCpre, anterior cingulate cortex, pregenual; ACCsup, anterior cingulate cortex, supracallosal; pIns, posterior insula; aIns, anterior insula; MNI, Montreal Neurological Institute. To localize the regions, we employed a probabilistic atlas for the insula cortex (Faillanot et al., 2017), and the Automated Anatomical Atlas (AAL3) (Rolls et al., 2020) for all the other regions.

##### Description of the selected studies

DeCharms et al. 2005 used rt-fMRI-NF to study how the willful modulation of MCC affects acute pain and chronic pain. Testing 8 healthy participants and 4 control groups under different conditions, they observed that only the experimental group learned to modulate the activity of the MCC and concomitantly reported a reduction in the level of perceived acute pain. In the same study, the Authors investigated 12 subjects (8 experimental and 4 control) with chronic pain observing that the regulation of MCC activity reduced the level of perceived chronic pain. Emmert et al. 2014 studied in two groups of healthy subjects (7 subjects per group) whether the willful modulation of subgenual ACC and aIns activity decreased the level of acute pain perception. Both groups reduced pain during the rt-fMRI-NF, but the Authors reported no difference in the level of pain between regulators (participants able to change the activity of the selected target) and non-regulators. Rance et al. 2014b showed that 10 healthy subjects learned to downregulate left pIns and subgenual ACC and upregulate left pIns consistently but not subgenual ACC. Despite the learned ability, successful regulation was not linked to a reduction of pain intensity. Similarly, in their second study (Rance et al. 2014a), they trained participants to increase the difference in subgenual ACC and left pIns activity, but again, they did not find significant changes in pain perception. Guan et al. 2015, in a double-blinded randomized study, showed that 6 out of 8 post-herpetic neuralgia patients learned to regulate pregenual ACC activity with significant pain reduction.

#### 3.1.2. Functional surgery records selection

Details of the selected papers are reported in Table 3, and the corresponding identified coordinates for the targets are reported in Table 4. The coordinates calculated by reviewers 2 (LM) and 3 (AN) were less than 5 mm from the coordinate of reviewer 1 (SF) (mean: 2.5 mm, range: 0-5 mm).

**Table 3:** Functional Neurosurgery studies.

| Author | Target | Target regions identified | Population | Sample | Technique | Stimulation effects on pain |
| --- | --- | --- | --- | --- | --- | --- |
| Rasche et al. (2006) | VPL/VPM | "The coordinate for the lateral somatosensory thalamus was as follows: Y = 3 to 5 mm anterior to the posterior commissure; Z = 0 to -2 mm below the inter commissural line; and X = 10 to 12 mm lateral to the midline for facial pain, 12 to 15 mm for pain in the upper extremity, and 15 to 18 mm for pain in the lower extremity. The coordinates for PVG were as follows: Y = 2 to 3 mm anterior to the posterior commissure; Z = 2 mm above to 2 mm below the intercommissural line; and X = 2 mm lateral to the wall of the third ventricle." | chronic neuropathic, mixed nociceptive/neuropathic pain | 56 | DBS of thalamic nuclei | "The best long-term results were seen in patients with failed-Back Surgery Syndrome. Patients with peripheral neuropathic pain and Dysesthesia Dolorosa responded very well to DBS. The results in Spinal Cord Injury patients with central pain syndromes were less favorable. Disappointing outcomes for patients with thalamic pain syndrome or post-stroke pain." |
| Boccard et al. (2017) | MCC | "The target used was 20mm posterior to the anterior tip of the frontal horns of the lateral ventricles." | chronic neuropathic pain | 24 | DBS in cingulate cortex | "The NRS dropped significantly." |
| Abreu et al. (2017) | VPL/VPM | VPL found 10–13 mm lateral to the posterior commissure | chronic neuropathic pain | 16 | DBS of thalamic nuclei | "Patient-reported outcome measure improvements were clear after three years in this cohort. The VAS, UWNPS, and BPI scores have improved with statistical significance after one month. Throughout the third year, mean pain relief was sustained." |
| Abdallat et al. (2021) | CM/Pf<br>VPL/VPM | "The CM/Pf targets were x = 8 mm lateral to the inter-commissural line, y = 8 mm posterior to the mid-commissural point, and z = 0 at the level of the inter-commissural plane. Target coordinates for the VPL were: x = 14, y = -10, and z = 0; and for the VPM: x = 12, y = -10, and z = 0" | chronic neuropathic pain | 40 | DBS of thalamic nuclei | "Out of the total group of 40 patients, 33 (82.5%) indicated that they had a clear benefit during test stimulation." |
| Strauss et al. (2018) | MCC | "The target on each side, 24 mm behind the tip of the frontal horn, 7–8 mm from the midline the second lesion was performed more anteriorly, 16 mm behind the tip of the frontal horn." | intractable oncological pain | 13 | Surgical cingulotomy | "All patients reported significant pain relief immediately after the operation in the recovery room (mean VAS 0.9 ± 1.3). Eight of the 11 patients (72.7%) reported significant pain relief (<50%), and 1 patient (9%) reported partial improvement. In 2 patients, the pain returned to its original severity." |
| Yen et al. (2005) | MCC | "The target slice was selected as 24 mm posterior to the anterior tip of the lateral ventricle." | intractable oncological pain | 22 | Surgical cingulotomy | "Of the 15 patients with cancer pain, significant or meaningful pain relief was achieved in 67% of patients at one-month follow-up, which decreased to 58% at three months and 50% at six months. Of the seven patients with intractable pain from non-neoplastic origin, four achieved significant pain relief, one obtained meaningful relief, and two reported no change at one-year follow-up." |
| Gallay et al. (2023) | CLp | "This corresponds to the anteroposterior position of the CLp, centered between 3 mm anterior and 1 mm posterior to the posterior commissure." | chronic neuropathic pain | 55 | MRgFUS thalamotomy | "The mean pain relief rated by patients was 42% ± 32% at 3 months, 43% ± 36% at 1 year, and 42% ± 37% at the last follow-up (n = 63)" |
| Urgosik et al. (2018) | CM/pf | "The CM/Pf complex was localized 4–6 mm lateral to the wall of the third ventricle, 8 mm posterior to the midpoint, and 2–3 mm superior to the inter commissural line" | chronic trigeminal neuralgia | 30 | Gamma knife thalamotomy | "Initial successful results were achieved in 13 (43.3%) of the patients, with complete pain relief in 1 of these patients." |
| Lovo et al. (2019) | CM/pf | "The best coordinates of CM/pf were X: 5.5 mm from the thalamic border, Y: 3.7 mm anterior to the posterior commissure, and Z: 3.7 mm from the inter commissural line." | refractory trigeminal pain<br>other complex pain syndromes | 14 | Gamma knife thalamotomy | "90% patients reported some form of relief, the average VAS at the time of response was 3.5(range: 0-9), and the average time to response was 67.3 days (range: 2-210). The neuromodulation effect of radiation was seen in 60% of patients." |

Abbreviations: MCC, middle cingulate cortex; CLp, posterior section of the central lateral thalamus; CM/Pf, centromedian/parafascicular nucleus; VPL, ventral posterior lateral nucleus; VPM, ventral posterior medial nucleus; DBS, Deep Brain Stimulation; VAS, Visual Analog Scale; NRS, Numerical Rating Scale.

**Table 4:** Coordinates of the targets of the functional neurosurgery studies.

| Author | Region | Radius (mm) | MNI coordinates |  |  |  |  |  |
| --- | --- | --- | --- | --- | --- | --- | --- | --- |
|  |  |  | Left hemisphere |  |  | Right hemisphere |  |  |
| Yen et al. (2005) | MCC | 6 | -6 | 5 | 34 | 6 | 5 | 34 |
| Boccard et al. (2017) | MCC | 6 | -7 | 9 | 31 | 7 | 9 | 31 |
| Strauss et al. (2018) | MCC | 6 | -8 | 9 | 33 | 8 | 9 | 33 |
| Urgosik et al. (2018) | CM/pf | 3 | -8 | -19 | 0 | 8 | -19 | 0 |
| Lovo et al. (2019) | CM/pf | 3 | -8 | -22 | 3 | 8 | -22 | 3 |
| Abdallat et al. (2021) | CM/pf | 3 | -8 | -20 | -1 | 8 | -20 | -1 |
| Rasche et al. (2006) | VPL/VPM | 3 | -19 | -21 | 6 | 18 | -20 | 6 |
| Abreu et al. (2017) | VPL/VPM | 3 | -19 | -21 | 6 | 18 | -20 | 6 |
| Abdallat et al. (2021) | VPL/VPM | 3 | -19 | -21 | 6 | 18 | -20 | 6 |
| Gallay et al. (2023) | CLp | 3 | -11 | -25 | 6 | 11 | -25 | 6 |

Abbreviations: funcSurg, functional neurosurgery studies; MCC, middle cingulate cortex; CLp, posterior section of the central lateral thalamus; CM/Pf, centromedian/parafascicular nucleus of the thalamus; VPM/VPL, ventral posterior medial and ventral posterior lateral nucleus of the thalamus; MNI, Montreal Neurological Institute.

##### Deep brain stimulation (DBS)

Based on the results of the meta-analyses selection (see Figure 2), for DBS, we capitalized on the recent work of Frizon et al. 2020, which identified 12 studies. From these papers, 3 were excluded because of the sample size (Hollingworth et al. 2017; Hunsche et al. 2013; Lempka et al. 2017), 2 since they did not sufficiently characterize the target (Coffey 2001; Yamamoto et al. 2006), and one (Hamani et al. 2006) because no successful effect on pain was found. Of the remaining 6 studies, 3 from the same group (Abreu et al. 2017; Boccard et al. 2013; Pereira et al. 2013) reported the same localization of the target in the thalamic nuclei, the other 2 papers also from the same group (Boccard et al. 2014, 2017) localized the target in the cingulate cortex. In agreement with the AAL3 atlas, this region was the MCC, although the Authors referred to it as the anterior cingulate cortex, in accordance with the different nomenclature. For all these papers reporting the same target location, the target was identified through the description found in Abreu et al. 2017 for VPL/VPM and in Boccard et al. 2017 for the MCC. Thus, from the remaining 3 papers, we extracted the localization of targets located in VPL/VPM (Abreu et al. 2017; Rasche et al. 2006), and in the cingulate cortex (Boccard et al. 2017). The search for more recent papers (see Figure 3) with the selected criteria identified Abdallat et al. 2021 targeting the centromedian/parafascicular nucleus (CM/Pf) and VPL/VPM and Abreu et al. 2022 which, however, reported a follow-up of a previous study and thus was excluded. Since the computed MNI coordinates of VPM/VPL in Abdallat et al. 2021, Abreu et al. 2022, and Rasche et al. 2006 did not appear to be at the expected position when superimposed on the thalamic atlas in the MNI space (Saranathan et al. 2021), we relied on the center of mass of VPL/VPM extracted from the same atlas. Notably, we selected only one paper, also reporting coordinates of VPL/VPM, reporting the coordinates of the PAG/PVG (Rasche et al. 2006). However, since the extracted PAG/PVG coordinates appeared not to be in the expected position, they were not selected for the successive analyses.

**Figure 2:**
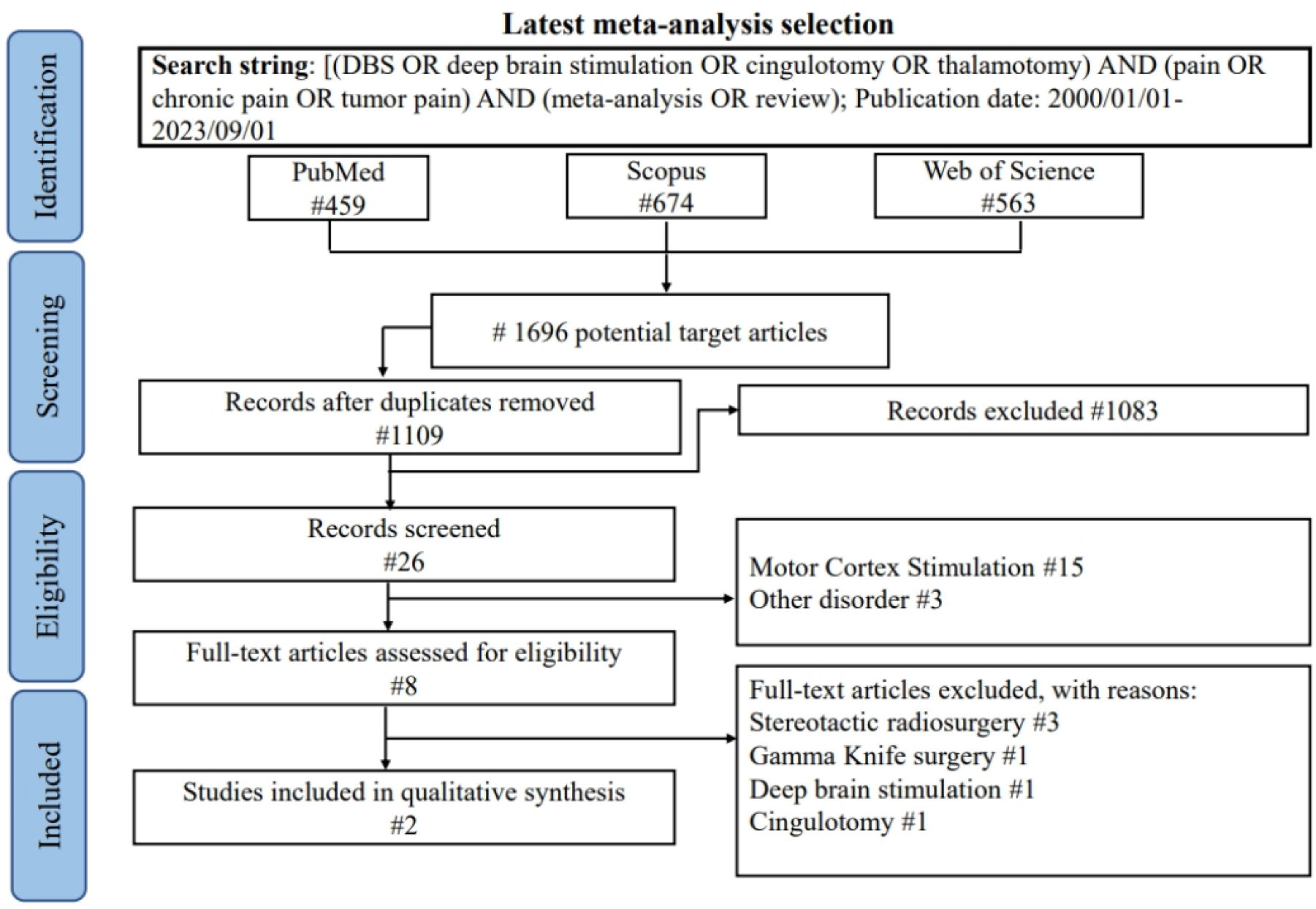
Flowchart of the screening process of the funcSurg meta-analyses according to the PRISMA guideline.

**Figure 3:**
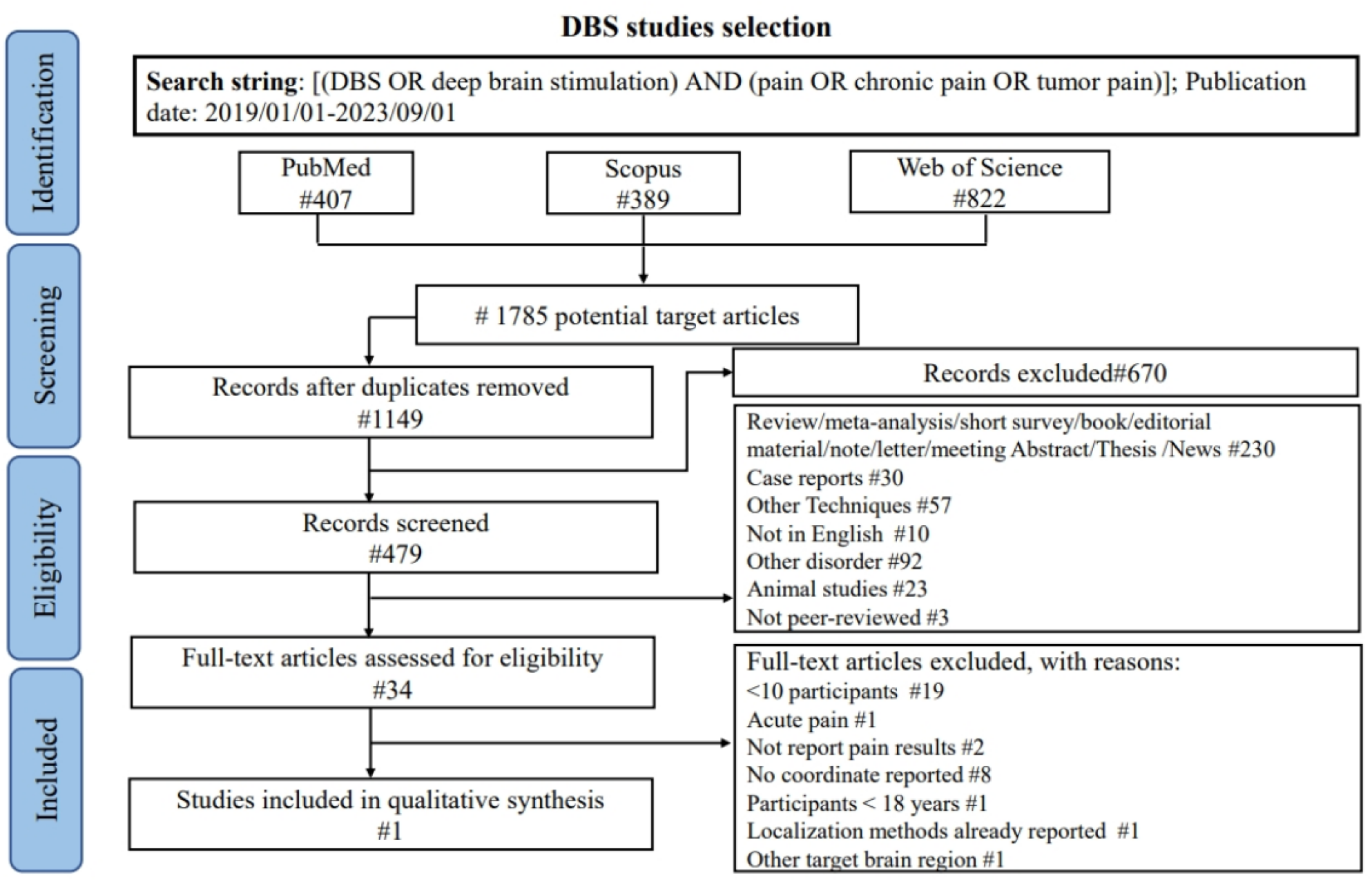
Flowchart of the screening process of the deep brain stimulation (DBS) studies according to the PRISMA guideline.

##### Cingulotomy

In the selected review (Sharim and Pouratian 2016), 11 studies were reported; however, 8 were published before 2000. This left 2 studies from the same group (Yen et al. 2005, 2009) reporting the same target localization and 1 study (Patel et al. 2015) on 3 patients that was therefore discarded. We also searched for more recent papers from January 2016 to August 2023 (see Figure 4) and we found a study by Wang et al. 2017 employing the same target localization as reported by Yen et al. 2005, the work from Strauss et al. 2018 investigating a slightly different localization of the target, and the work from Hochberg et al. 2020 using the same target localization as reported by Strauss et al. 2018. So, for the localization of the cingulate target, we relied on the papers of Yen et al. 2005 and Strauss et al. 2018.

**Figure 4:**
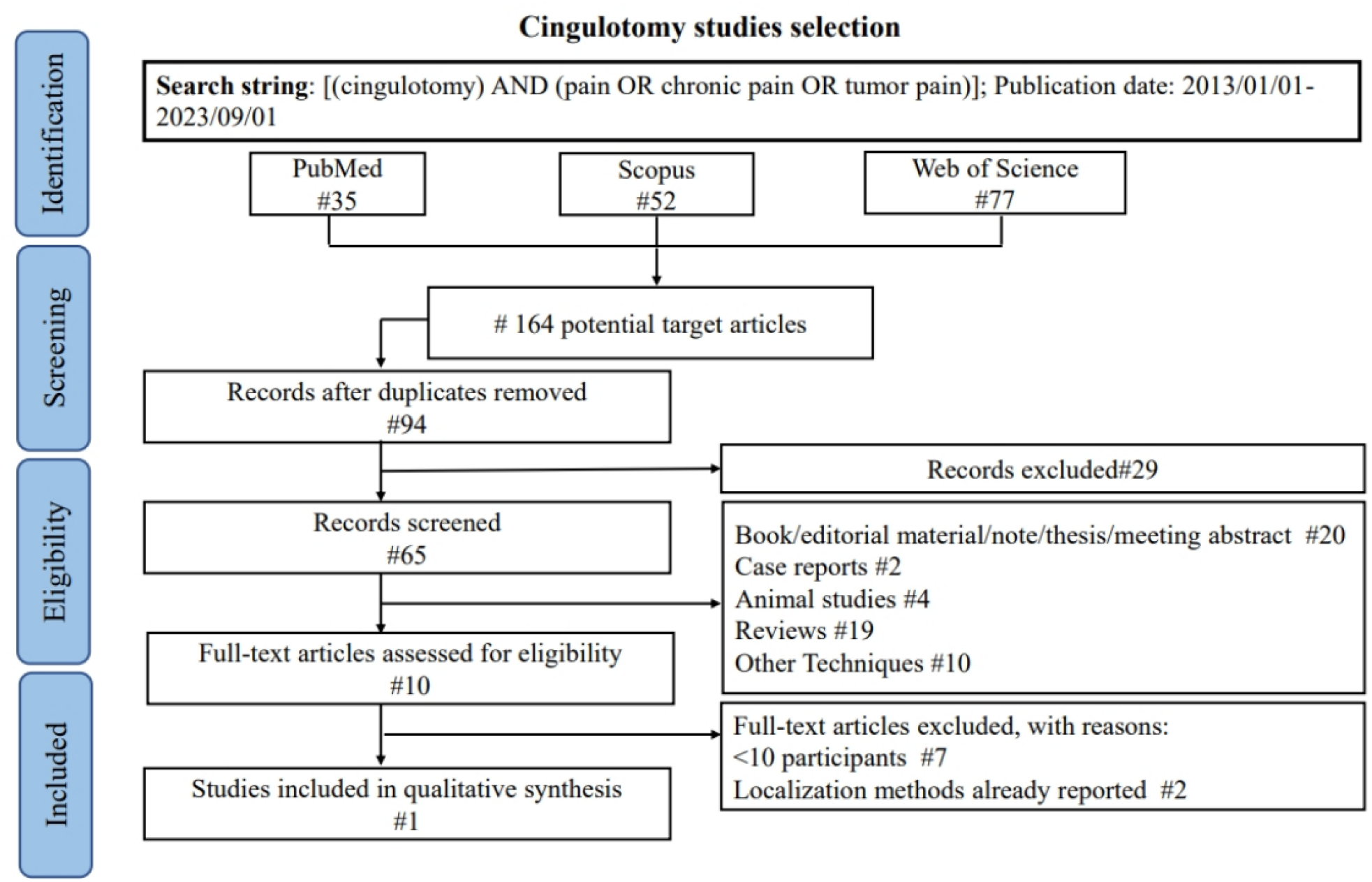
Flowchart of the screening process of the cingulotomy studies according to the PRISMA guideline.

##### Thalamotomy

The literature search (see Figure 5), according to the selected criteria, identified 5 papers targeting the posterior central lateral thalamic nuclei (CLp) (Gallay, Moser, and Jeanmonod 2020; Gallay et al. 2023; Jeanmonod et al. 2001; Jeanmonod et al. 2012) and the CM/Pf complex (Lovo et al. 2019; Urgosik and Liscak 2018). Notably, for the CLp, several findings were reported by the same group (Gallay et al. 2023; Jeanmonod et al. 2001; Jeanmonod et al. 2012) with the target being updated in their most recent paper. (Gallay et al. 2023). Thus, we considered only the most recent study (Gallay et al. 2023).

**Figure 5:**
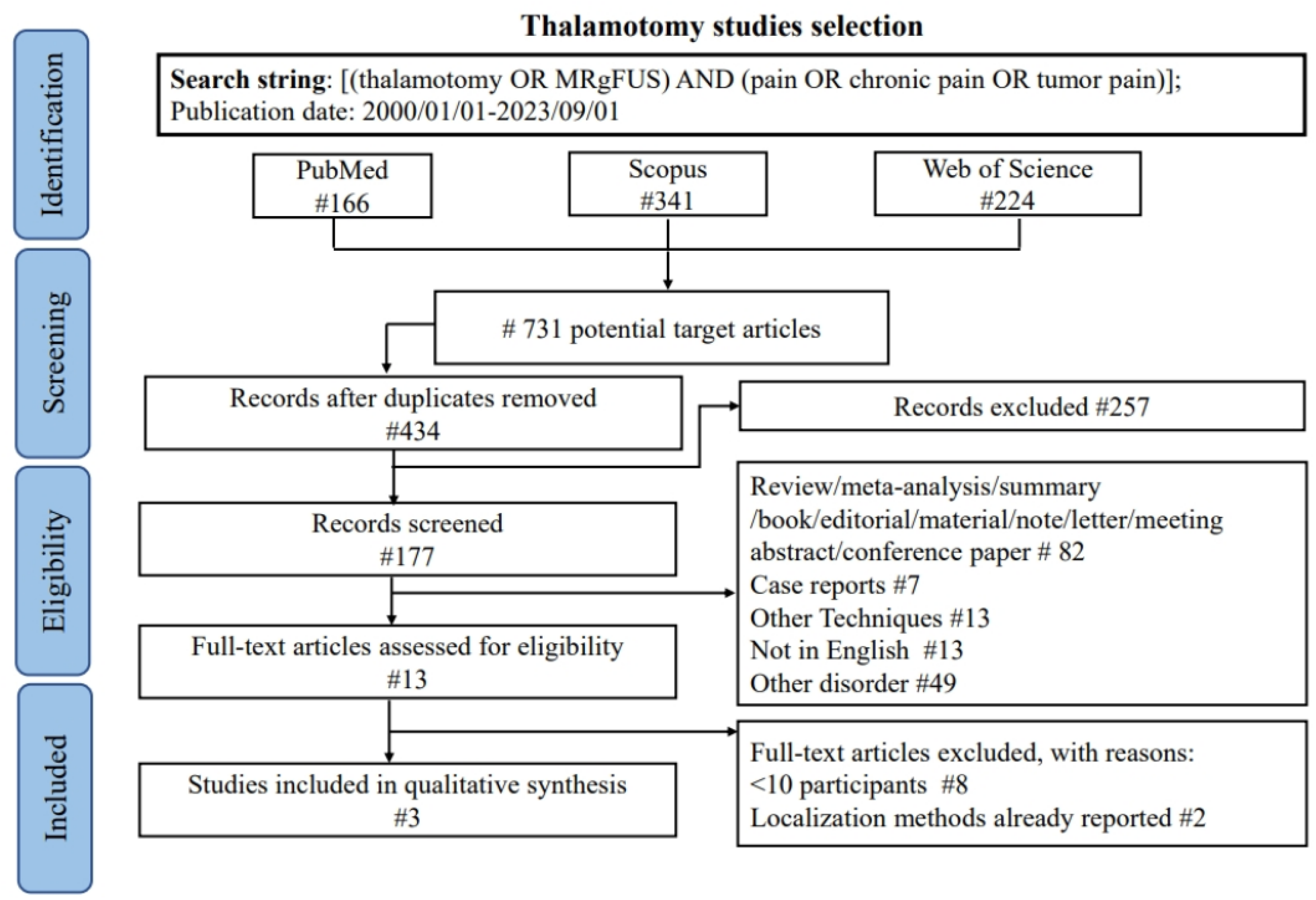
Flowchart of the screening process of the thalamotomy studies according to the PRISMA guideline.

### 3.2. Identification of the rs-fMRI networks underlying the target regions

Based on the identified MNI coordinates, 8 ROIs for the rt-fMRI-NF records (8 targets identified in right MCC, left aIns, left pIns, and left pregenual, subgenual, and supracallosal ACC) and 8 bilateral ROIs [targets identified in CLp, CM/Pf, VPL/VPM, and MCC] for funcSurg records were built to produce functional connectivity maps (or SBC maps) on the 7T MRI HCP dataset. For a detailed report of the identified rs-fMRI networks, see Figure 6.

**Figure 6:**
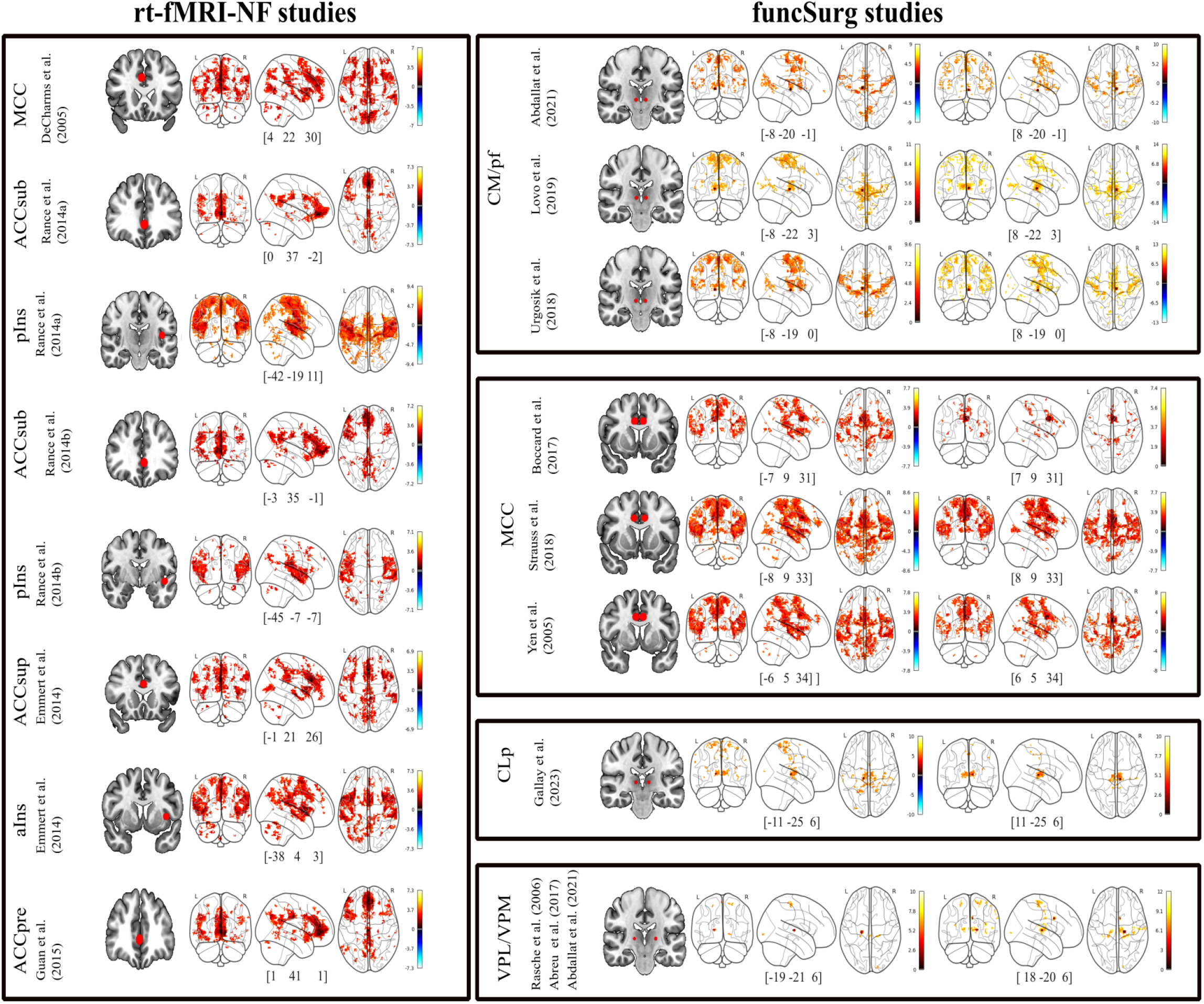
Seed-based rs-fMRI maps (obtained from a subsample of the 7T MRI Human Connectome Project dataset) of each identified target in the selected rt-fMRI-NF and funcSurg studies. Data were considered significant for cluster threshold: p<0.05 cluster-level, p-FDR corrected and voxel threshold: p <0.001 uncorrected. Abbreviations: rt-fMRI-NF, real-time fMRI neurofeedback; funcSurg: functional neurosurgery; MCC, middle cingulate cortex; ACC, anterior cingulate cortex (ACCsub, subgenual; ACCsup, supracallosal; ACCpre, pregenual); pIns, posterior insula; aIns, anterior insula; CM/Pf, centromedian/parafascicular nucleus of the thalamus; CLp, posterior section of the central lateral thalamus; VPL/VPM, ventral posterior lateral nucleus/ventral posterior medial nucleus of thalamus.

### 3.3. Principal components analysis of the identified rs-fMRI networks (PCA rs-fMRI-derived maps)

The PCA applied to the HCP rs-fMRI maps obtained from the rt-fMRI-NF targets (see Figure 7) identified 2 components, accounting for a cumulative explained variance of 84.7%. The component 1 accounted for 76% of the total variance, indicating that a substantial portion of the neural activity across the rt-fMRI-NF ROI rs-fMRI HCP maps is captured by a single neural network. This bilateral network includes the ACC, comprising the rostral/subgenual regions, the posterior cingulate cortex, and the antIns. The component 2, accounting for 8.7% of the total variance, showed prominent bilateral clusters in the MCC, in sensorimotor areas (precentral and postcentral areas, paracentral lobule, and supplementary motor area), and the insula cortex (in both postIns and antIns). Regarding, the PCA performed on the HCP rs-fMRI maps from the funcSurg targets (see Figure 7) revealed 4 components with a cumulative explained variance of 83%. The component 1, explaining 37% of the variance, encompasses a bilateral network comprising the insula cortex (antIns and postIns), the MCC, and the sensorimotor areas (precentral and postcentral areas, paracentral lobule, and supplementary motor area). The component 2, accounting for 20.4% of the variance, was characterized by a bilateral network comprising the MCC, parietal regions (comprising the superior parietal areas and the supramarginal gyrus), and areas of the insula cortex (mainly postIns). The component 3, accounting for 13.6% of the total explained variance, comprised mainly bilateral sensorimotor areas (precentral and postcentral areas, paracentral lobule, and supplementary motor area) and MCC with large clusters in the thalamus and the visual areas. The component 4 (12% of the total explained variance), was characterized by a bilateral network comprising the sensorimotor areas (precentral and postcentral areas, paracentral lobule, and supplementary motor area), the MCC, and clusters in the insula cortex covering mainly the posterior regions.

**Figure 7:**
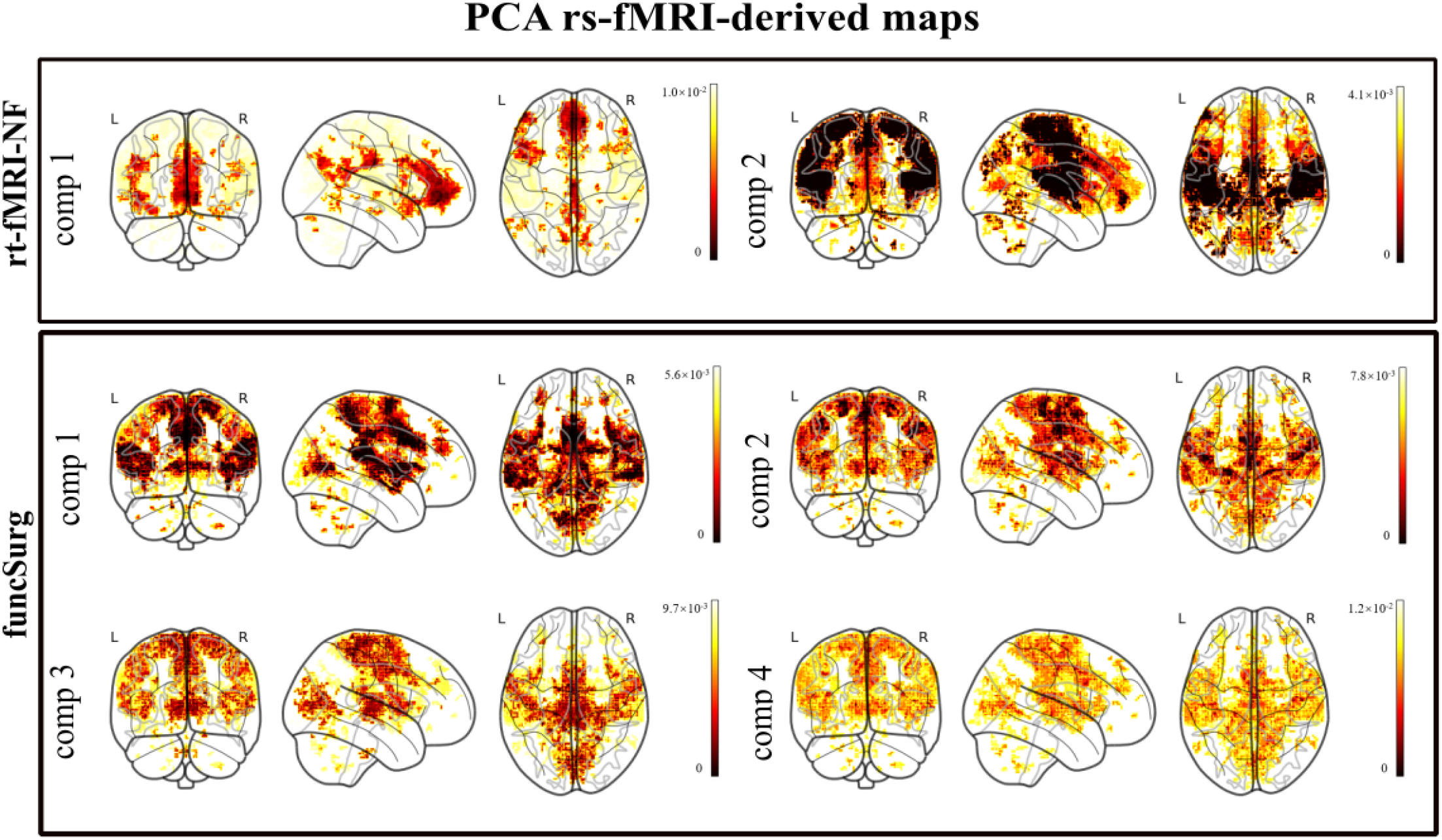
PCA rs-fMRI-derived maps. Abbreviations: rt-fMRI-NF, real-time fMRI neurofeedback studies; funcSurg, functional neurosurgery studies; comp, PCA component

### 3.4. Neurosynth decoding of PCA rs-fMRI-derived maps

In the Neurosynth decoding analyses (see Figure 8), component 1 of the rt-fMRI-NF PCA rs-fMRI-derived map correlated with functional terms related to self-referential processing (‘referential’, ‘self-referential’), reward processing (‘money’, ‘reward’, ‘value’) and with anatomical terms referring to the DMN (e.g., ‘default mode’, ‘pcc’, ‘network dmn’, ‘vmpfc, ‘mpfc’) and the anterior cingulate cortex (‘anterior cingulate’, ‘cortex acc’, ‘acc’). Component 2 presented correlations mainly with functional and anatomical terms related to sensorimotor processing (‘stimulation’, ‘motor’, ‘speech production’, ‘production, ‘tactile’) and network (e.g., ‘somatosensory’, ‘sensorimotor’, ‘posterior insula’, ‘SII’, ‘S1’) with terms specifically related to the motor cortex (‘motor cortex’, ‘primary motor’, ‘motor cortex’).

**Figure 8:**
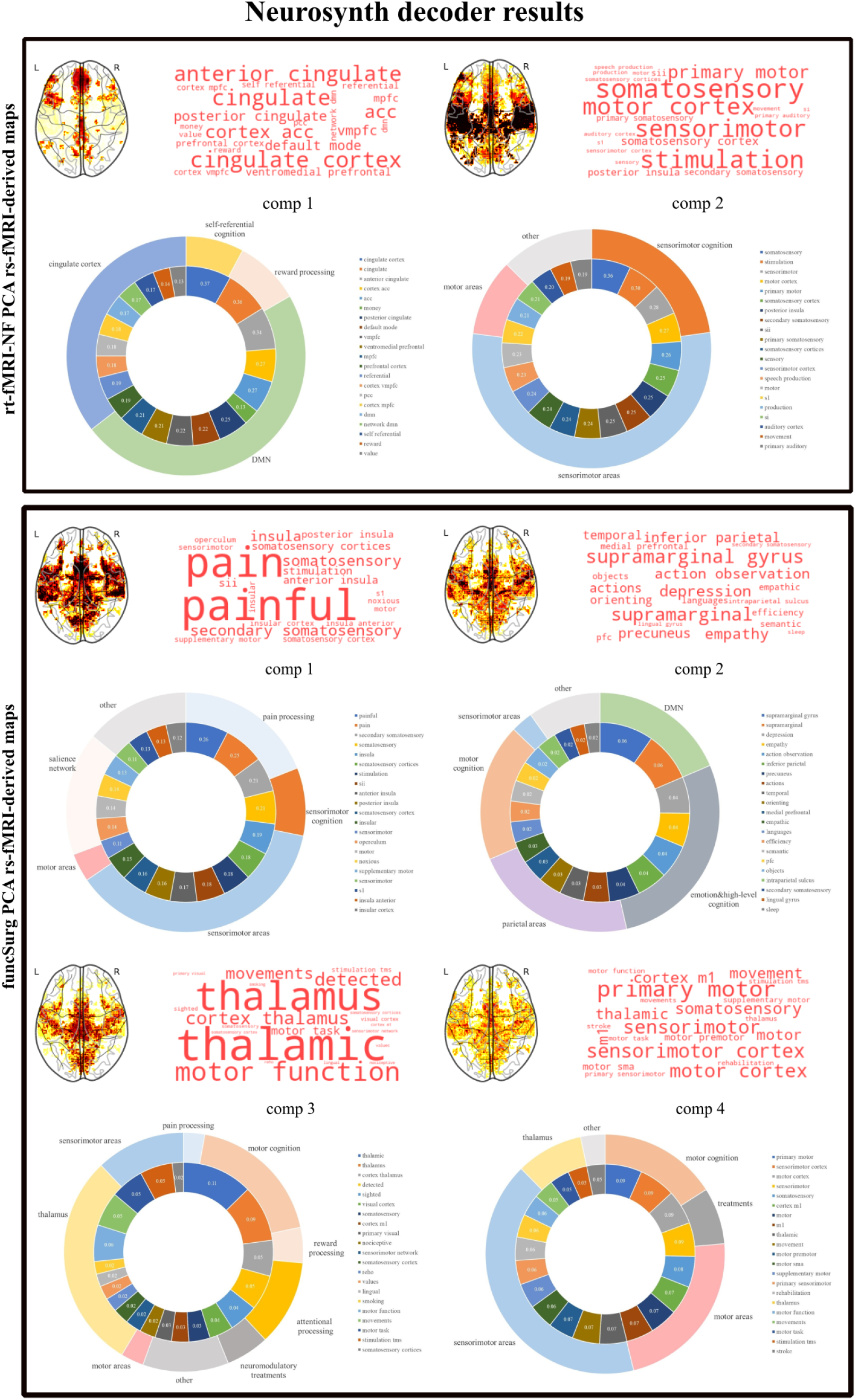
Neurosynth decoding of PCA rs-fMRI-derived maps retaining the top 20 terms with their correlation values in the inner circle. Abbreviations: rt-fMRI NF, real-time fMRI neurofeedback studies; funcSurg, functional surgery studies; PCA, principal component analysis; comp, component.

Regarding the funcSurg PCA rs-fMRI-derived maps, for component 1, functional terms were referred to pain (i.e., ‘pain,’ ‘painful,’ ‘noxious’) and sensorimotor cognition (e.g., ‘motor,’ ‘stimulation’). Anatomical terms were referred to the sensorimotor network (e.g., ‘somatosensory,’ ‘S1’, ‘SII,’ ‘supplementary motor,’ ‘posterior insula’), but also to the salience network (e.g., ‘anterior insula’). For component 2, functional terms were related to sensorimotor cognition (e.g., ‘motor’ and ‘actions’,’orienting’, ‘action observation’, ‘actions’, ‘objects’) and to terms related to emotions and high-level cognition (e.g., ‘depression’, ‘empathy’, ‘languages’, ‘semantic’, ‘efficiency’), while anatomical terms were referred to the parietal regions (e.g., ‘supramarginal gyrus’, ‘inferior parietal’, ‘intraparietal sulcus’), and to regions of the DMN (e.g., ‘precuneus’, ‘medial prefrontal’, ‘inferior frontal’) and the sensorimotor areas (e.g., ‘secondary somatosensory’). For component 3, functional terms were referred to pain (e.g., ‘nociceptive’) and sensorimotor cognition (e.g., ‘motor task’, ‘movement’), but also included terms related to reward (e.g., ‘values’, ‘smoking’) and attentional processing (e.g., ‘detected’, ‘sighted’), and neuromodulatory treatments (e.g., ‘stimulation tms’). Anatomical terms were mainly related to the sensorimotor areas (e.g., ‘somatosensory’, ‘cortex m1’), but also included the visual cortex (e.g., ‘visual cortex’, ‘primary visual’) and the thalamus (e.g., ‘thalamic’, ‘thalamus’). For Component 4, functional terms were related to sensorimotor cognition (e.g., ‘movements,’ ‘motor function,’ ‘motor task’), as well as to treatments (e.g., ‘stimulation tms,’ ‘rehabilitation’). Similarly, anatomical terms were related to the sensorimotor areas (e.g., ‘sensorimotor cortex,’ ‘primary sensorimotor,’ ‘sensorimotor’), with several terms specifically related to the motor cortex (e.g., ‘motor sma,’ ‘supplementary motor’, ‘m1’).

### 3.5. Identification of membership of PCA rs-fMRI-derived maps in the canonical rs-fMRI networks

Formal tests of the similarity of the PCA rs-fMRI-derived maps to the canonical rs-fMRI networks (DMN, salience network, and sensorimotor network) showed that the rt-fMRI-NF component 1 and component 2 significantly overlapped with the DMN (respectively, 14.9% and 9.8% of shared voxels). Additionally, component 1 overlapped, although modestly, with the salience network (4% of shared voxels), while component 2 with the sensorimotor network (28.9% of shared voxels). Interestingly, all the selected funcSurg components were significantly similar to the salience network (with 41.7%, 14.2%, 14.8%, and 15% of shared voxels, respectively). Additionally, components 1, 3, and 4 also significantly overlapped with the sensorimotor network (respectively with 12.7%, 13.8%, and 20.4% of shared voxels, respectively) (see Table 5).

**Table 5:** Testing the similarity between PCA-derived rs-fMRI maps and canonical rs-fMRI networks (salience network, DMN, and sensorimotor network)

| Maps | SN |  | DMN |  | SMN |  |
| --- | --- | --- | --- | --- | --- | --- |
|  | *p*-value | *emp* | *p*-value | *emp* | *p*-value | *emp* |
| rt-fMRI-NF PCA map1 | 0.006* | 0.040 | 0.001* | 0.149 | 0.999 | 0.000 |
| rt-fMRI-NF PCA map2 | 0.103 | 0.078 | 0.002* | 0.098 | 0.001* | 0.289 |
| funcSurg PCA map1 | 0.001* | 0.417 | 0.964 | 0.054 | 0.023* | 0.127 |
| funcSurg PCA map2 | 0.001* | 0.142 | 0.874 | 0.023 | 0.946 | 0.016 |
| funcSurg PCA map3 | 0.001* | 0.148 | 1.000 | 0.156 | 0.001* | 0.138 |
| funcSurg PCA map4 | 0.001* | 0.150 | 0.999 | 0.026 | 0.001* | 0.204 |
| NS: term 'chronic pain' | 0.001* | 0.105 | 0.995 | 0.008 | 0.997 | 0.003 |
| NS: term 'pain' | 0.001* | 0.490 | 0.928 | 0.069 | 0.994 | 0.037 |

Abbreviations: *emp*, empirical value; SN, Salience Network; DMN, Default Mode Network; SMN, Sensorimotor Network; rt-fMRI-NF PCA: PCA rs-fMRI-derived map of real-time fMRI neurofeedback targets; PCA funcSurg PCA: PCA rs-fMRI-derived map of functional surgery targets, NS: Neurosynth meta-analytic map; \* *p*-value <0.05.

### 3.6. Neurosynth meta-analytic brain maps of ‘pain’ and ‘chronic pain terms’

Uniformity maps obtained from Neurosynth showed that the term “chronic pain” was associated with robust activity in clusters centered in the bilateral insula, inferior parietal cortex, and amygdala. These clusters extended to the MCC, posterior regions of the ACC, supplementary motor area, and subcortical regions such as the hippocampus and thalamic nuclei. The more extensive “pain” uniformity map showed that this term was mainly associated with activity in the bilateral insula, pregenual cingulate cortex and subgenual regions of the frontal areas, motor areas (supplementary and pre-supplementary), including the MCC, parietal regions, and amygdala (for details). The formal test employing null maps with preserved spatial autocorrelation showed that these two maps significantly overlapped with the salience network (10.5% overlapping voxels for the term ‘chronic pain’ and 49% for ‘pain’), but not with the DMN nor the sensorimotor network (see Table 5).

### 3.7. Neurotransmitter receptors profiling of PCA maps

Spatial correlation analysis between each PCA rs-fMRI-derived map and the neurotransmitter receptor distribution maps showed that, in the case of the rt-fMRI-NF targets, component 1 was significantly correlated with the FDOPA(f18) receptor map, while component 2 with the NAT receptor map. Regarding funcSurg targets, spatial correlation analysis revealed consistent patterns of significant correlations (3 out of 4 components) with the NAT receptor map (see Figure 9 and Table 6 for details).

**Table 6:** Neurotransmitter receptors profiling of the PCA rs-fMRI-derived maps.

| PET Map | funcSurg<br>PCA comp1<br>p-value | funcSurg<br>PCA comp2<br>p-value | funcSurg<br>PCA comp3<br>p-value | funcSurg<br>PCA comp4<br>p-value | rt-fMRI-NF<br>PCA comp1<br>p-value | rt-fMRI-NF<br>PCA comp2<br>p-value |
| --- | --- | --- | --- | --- | --- | --- |
| 5HT1a(WAY) | 0.018 | 0.571 | 0.760 | 0.905 | 0.686 | 0.433 |
| 5HT1a(cumi) | 0.140 | 0.459 | 0.732 | 0.857 | 0.713 | 0.884 |
| 5HT1b(P943) | 0.001** | 0.898 | 0.061 | 0.065 | 0.111 | 0.379 |
| 5HT1b(az) | 0.060 | 0.611 | 0.844 | 0.356 | 0.183 | 0.456 |
| 5HT2a(ALT) | 0.001** | 0.683 | 0.169 | 0.094 | 0.060 | 0.739 |
| 5HT2a(cimbi) | 0.010 | 0.464 | 0.599 | 0.173 | 0.116 | 0.795 |
| 5HT4(sb20) | 0.485 | 0.893 | 0.835 | 0.931 | 0.627 | 0.819 |
| CB1(PFMPEPd2) | 0.001** | 0.308 | 0.853 | 0.416 | 0.023 | 0.849 |
| D1(SCH23390) | 0.326 | 0.556 | 0.201 | 0.457 | 0.057 | 0.516 |
| D2(RACLOPRIDE) | 0.141 | 0.350 | 0.219 | 0.060 | 0.321 | 0.232 |
| D2(fallypride) | 0.615 | 0.562 | 0.831 | 0.721 | 0.567 | 0.212 |
| DAT(DATSPECT) | 0.092 | 0.114 | 0.953 | 0.918 | 0.007 | 0.530 |
| FDOPA(f18) | 0.090 | 0.128 | 0.927 | 0.823 | 0.001** | 0.473 |
| GABAa(FLUMAZENIL) | 0.001** | 0.386 | 0.112 | 0.015 | 0.141 | 0.475 |
| GABAa(flumazenil) | 0.051 | 0.974 | 0.776 | 0.205 | 0.234 | 0.656 |
| KappaOp(LY2795050) | 0.001** | 0.518 | 0.562 | 0.206 | 0.018 | 0.205 |
| MU(CARFENTANIL) | 0.952 | 0.464 | 0.136 | 0.763 | 0.471 | 0.950 |
| MU(carfentanil) | 0.452 | 0.659 | 0.280 | 0.835 | 0.864 | 0.862 |
| NAT(MRB) | 0.001** | 0.001** | 0.323 | 0.001** | 0.019 | 0.001** |
| NMDA(ge179) | 0.740 | 0.041 | 0.156 | 0.046 | 0.551 | 0.211 |
| SERT(DASB) | 0.572 | 0.043 | 0.155 | 0.775 | 0.002 | 0.946 |
| SERT(MADAM) | 0.823 | 0.130 | 0.187 | 0.529 | 0.034 | 0.744 |
| SERT(dasb) | 0.269 | 0.082 | 0.611 | 0.691 | 0.015 | 0.858 |
| VAcHT(feobv1) | 0.586 | 0.028 | 0.243 | 0.526 | 0.437 | 0.003 |
| VAcHT(feobv2) | 0.111 | 0.012 | 0.582 | 0.899 | 0.005 | 0.027 |
| VAcHT(feobv3) | 0.001** | 0.086 | 0.193 | 0.456 | 0.027 | 0.056 |
| mGluR5(abp1) | 0.003 | 0.638 | 0.443 | 0.267 | 0.348 | 0.471 |
| mGluR5(abp2) | 0.001** | 0.963 | 0.284 | 0.185 | 0.525 | 0.353 |
| mGluR5(abp3) | 0.001** | 0.968 | 0.392 | 0.115 | 0.271 | 0.378 |

\*\* Results were considered significant for $p$ -value $<0.0017$ - Bonferroni corrected for 29 statistical tests.
Abbreviations: funcSurg, functional neurosurgery; rt-fMRI-NF, real-time functional magnetic resonance imaging neurofeedback; PCA, principal component analysis; comp, component. For an extended explanation of the neurotransmitter maps employed, see Dukart et al. (2020).

**Figure 9:**
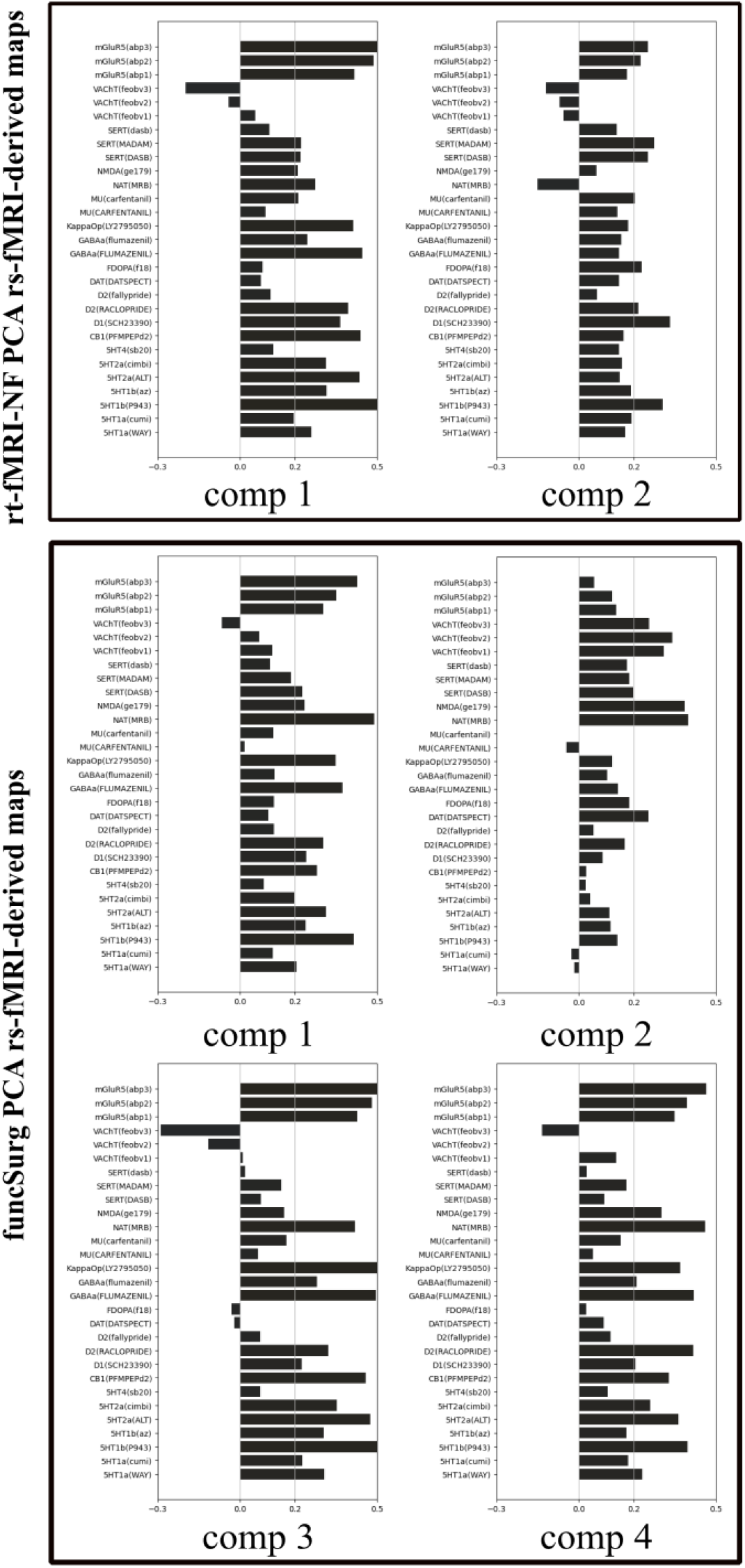
Correlation coefficients between PCA rs-fMRI-derived maps and neurotransmitter receptor distribution maps available in Juspace. Results were considered significant for p-value<0.0017, Bonferroni correction for 29 statistical tests to control for multiple comparisons. For an extended explanation of the neurotransmitter maps employed, see Dukart et al. (2020).

## 4. Discussion

FuncSurg approaches for treating chronic pain, while considered quite effective in a good number of patients, are not routinely applied in clinical practice mainly because of their invasiveness. However, they provide highly-specific and potentially causal manipulations for informing target regions for techniques involving noninvasive neuromodulation targeting chronic pain, including rt-fMRI-NF.

Despite the remarkable potential of rt-fMRI-NF in the treatment of pain results remained inconsistent. Our study aimed to characterize the rt-fMRI-NF brain targets used in the current literature and to identify new ones, taking advantage of funcSurg studies that have shown that modulation or lesioning of specific brain regions can effectively mitigate chronic pain. To facilitate a comprehensive overview, after identifying the brain targets of rt-fMRI-NF and funcSurg studies, we characterized, using several unbiased approaches, the underlying functional networks from neurofunctional, meta-analytic, and neurotransmitter perspectives.

We report 3 main findings. Firstly, and as expected, we showed that the targets employed by rt-fMRI-NF are markedly different from those used in funcSurg studies, except for the MCC, employed in several funcSurg studies considered here (Boccard et al. 2017; Strauss et al. 2018; Yen et al. 2005) and in only one rt-fMRI-NF study (DeCharms et al. 2005).

Secondly, we showed that rt-fMRI-NF targets are major hubs of a single functional circuit that explains more than 70% of the variance in the data maps and that, according to the large-scale meta-analytic ‘decoding’ using Neurosynth, mainly maps onto regions typically assigned to the DMN (component 1). This was confirmed by comparing the overlap of the target maps with rs-fMRI networks from an independent rs-fMRI dataset (Shirer et al. 2012) using spatial autocorrelation preserving null maps (Burt et al. 2020) that, however, also suggests a modest involvement of the salience network. In contrast, all funcSurg targets, although with some differences, are hubs of sensorimotor networks (components 1,3, and 4) and of regions involved in sensorimotor cognition (components 1,2,3, and 4), with motor areas being highly prevalent. Also, this finding was confirmed by comparing the overlap of the target maps with rs-fMRI networks from an independent rs-fMRI dataset (Shirer et al. 2012), although in this case, the results indicated a robust overlap with the salience network for all the PCA rs-fMRI-derived networks. Thirdly, we showed that, although the rs-fMRI networks associated with funcSurg targets differ in their association with neurotransmitter systems, they converge, except component 3, on the noradrenergic system.

The meta-analysis on rt-fMRI-NF in acute and chronic pain revealed only 5 studies that examined healthy individuals (DeCharms et al. 2005; Emmert et al. 2014; Guan et al. 2015; Rance et al. 2014a,b) and chronic pain patients (DeCharms et al. 2005; Guan et al. 2015). These studies targeted different areas of the ACC (rostral, subgenual, pregenual, and supracallosal ACC) (Emmert et al. 2014; Guan et al. 2015; Rance et al. 2014a,b), the MCC (DeCharms et al. 2005), and of the insular cortex (Emmert et al. 2014; Rance et al. 2014a,b). In general, participants were able to learn to modulate the target regions, but with conflicting results at the behavioral level. Studies involving healthy participants (Emmert et al. 2014; Rance et al. 2014a,b), with the exception of DeCharms et al. 2005, showed no significant behavioral changes in pain levels during acute pain infliction. In this regard, it is worth noting that although in Emmert et al. 2014 the modulation of the targets (left sup ACC and left aIns) resulted in a reduction in acute pain perception, the lack of a control condition and the fact that both regulators and non-regulators showed a similar behavioral response do not allow robust conclusions. Only DeCharms et al. 2005, targeting MCC, reported a reduction in acute pain in healthy participants and, in the same study, a decrease in the level of chronic pain in patients. However, these results were not replicated in a subsequent study of the same group (Guan et al. 2015). Importantly, in chronic pain conditions, Guan et al. 2015 showed, in a double-blind randomized study, that the rt-fMRI-NF targeting of the pregenual ACC induced a reduction of the level of pain in 6 out of 8 post-herpetic neuralgia patients.

The meta-analysis on funcSurg studies applied to chronic pain identified 9 records that demonstrated a reduction of chronic pain in at least 40% of patients (Abdallat et al. 2021; Boccard et al. 2017; Gallay et al. 2023; Lovo et al. 2019; Rasche et al. 2006; Strauss et al. 2018; Urgosik and Liscak 2018; Yen et al. 2005). From these investigations, 8 bilateral targets were identified, specifically in the MCC (Boccard et al. 2017; Strauss et al. 2018; Yen et al. 2005) and in selected thalamic nuclei, namely VPL/VPM (Abdallat et al. 2021; Abreu et al. 2017; Rasche et al. 2006), CM/pf (Abdallat et al. 2021; Lovo et al. 2019; Urgosik and Liscak 2018) and CLp (Gallay et al. 2023).

It is worth noting that both the rt-fMRI-NF studies in chronic pain and the funcSurg approaches mainly investigated the effects of these treatments in neuropathic pain, although, of course, the funcSurg studies have treated patients in very severe conditions. Interestingly, despite the similar diagnosis of the involved patients, the above observations show that the anatomical targets of the rt-fMRI-NF and funcSurg approaches are quite different. Indeed, while the rt-fMRI-NF studies favored only cortical targets (particularly in the cingulate and insular area), the selected funcSurg approaches favored the thalamic nuclei and a single cortical target (in the MCC). These differences clearly lie in the diverse foundations of these approaches.

In this regard, rt-fMRI-NF studies are rooted in the relatively recent fMRI investigations of acute pain, identifying their targets in areas that respond to nociceptive stimulation, particularly in areas primarily involved in processing the emotional and cognitive aspects of pain. Although this may be a pragmatic approach and may reflect the slippage of chronic pain (as opposed to acute pain) toward the known alterations in areas involved in emotion processing (Kuner and Kuner 2020; Serafini, Pryce, and Zachariou 2020), the effectiveness of this method in defining brain areas that can modulate chronic pain remains to be fully understood. Today, we have ample evidence indicating that chronic pain is different from acute pain (Martucci, Ng, and Mackey 2014) and represents a distinct nosological entity with important neuroplastic reorganizations(Serafini, Pryce, and Zachariou 2020). Furthermore, new possible treatments to reduce chronic pain should target antinociceptive networks (Price et al. 2018), which, however, are still not well-defined at the forebrain and midbrain levels.

Differently, functSurg approaches to treat chronic pain are deeply rooted in a long tradition of neurosurgical and neurophysiological studies (Jeanmonod and Morel 2009). The medial thalamus, in particular, plays a significant role in the neurosurgical treatment of chronic pain. This thalamic region, which comprises some of the included funcSurg targets (i.e., CLp and CM/pf), is conceptualized as part of the medial pain system (Bushnell, Čeko, and Low 2013; De Ridder and Vanneste 2016; Kulkarni et al. 2005; Price 2000) and due to its afferent (from the spinothalamic and spino-reticular thalamic tracts) and efferent (to the aIns and dorsal ACC/MCC) connections, mainly overlapping with the salience network, it is considered to be involved in the motivational-affective aspects of pain (De Ridder, Adhia, and Vanneste 2021). This funcSurg target has a long history, as early as 1911 Head and Holmes hypothesized that an overactive ventral-posterior thalamic nucleus (VP), due to a reduction in its inhibition by the thalamic-cortical-thalamic loops, would generate abnormal, amplified impulses in the medial thalamus, inducing central pain. Later, Sano 1977 enriched this notion, introducing the idea that the generation of abnormal signals from this thalamic nucleus would induce an amplified activity of the reverberating circuit between the medial and lateral thalamus. Based on this knowledge, since about 1950, the medial thalamus has been the subject of funcSurg approaches, with the explicit goal of controlling its disinhibition (Jeanmonod and Morel 2009). More recently, Jeanmonod’s group, searching for a medial thalamic area with better clinical outcomes, identified the CLp as a new possible target, being part of a large thalamo-cortical network involved in the sensory cognitive and affective-motivational aspects of pain (Jeanmonod, Magnin, and Morel 1996).

Slightly more recent is the interest in the VPL/VPM developed in the wake of the investigation of Mazars, Merienne, and Cioloca 1980, who, stimulating the somatosensory pathways, induced paresthesias with chronic pain relief in patients with deafferentation pain (Rasche et al. 2005). These sensory thalamic nuclei, VPL/VPM according to the traditional view of the pathways of pain processing, are part of the lateral pain pathway (De Ridder and Vanneste 2016; Kulkarni et al. 2005; Price 2000), which maps onto the somatosensory cortex extending to the parietal area, and which mainly processes the discriminative/sensory components of pain (Bushnell, Čeko, and Low 2013; Flor et al. 1995; Kulkarni et al. 2005).

As for the MCC first pioneered in 1948 (Whitty et al. 1952), the neurosurgical lesioning of this region (described as anterior cingulate cortex in these studies) has been performed to relieve intractable pain, particularly terminal cancer pain (Allam et al. 2022). The apparent effects of cingulotomy not so much on nociceptive pain but on the suffering associated with it set the stage for recent approaches to both lesion and modulation of this brain region. MCC is considered a critical structure processing the affective-cognitive components of pain. However, it is also among the areas most commonly reported to be active across paradigms probing several cognitive and emotional domains and has been involved in several domains related to chronic pain, including central autonomic activation (Ferraro et al. 2022b).

Although the targets of the rt-fMRI-NF and funcSurg approaches are different, except for the MCC, it is possible that they could be different hubs within the same functional networks. Therefore, we identified the intrinsic networks underlying these regions by analyzing a subset of the HCP 7T rs-fMRI dataset. To allow easier interpretation, we reduced the number of rs-fMRI networks obtained (using PCA) separately for the rt-fMRI-NF and funcSurg studies.

For the sake of clarity of the next paragraphs, it is important here to specify that to identify the networks and regions best suited to be targeted with rt-fMRI-NF studies, we used two methods. The first method is based on a large-scale meta-analytic approach to perform quantitative functional and anatomical annotation, testing the association between terms present in the Neurosynth database with the obtained PCA-rs-fMRI-derived maps. The second method instead identifies the overlap and the associated p-value between the PCA rs-fMRI-derived maps and the relevant canonical rs-fMRI networks (obtained from an independent rs-fMRI dataset) using null maps that preserve the spatial autocorrelation of the original maps (Burt et al. 2020; Markello et al. 2021). From a broad perspective, we considered the approach employing the canonical rs-fMRI networks from a specific and limited dataset, as a lenient approach in comparison to the large-scale meta-analytic decoding obtained with Neurosynth.

Our results for the rs-fMRI-NF targets showed that only 2 components explained most of the variance of the underlying functional networks. Notably, the first component alone explained more than 74% of the variance, indicating that most targets mainly probe a single extended functional network that, as demonstrated by large-scale meta-analytic decoding with Neurosynth (Yarkoni et al. 2011), maps mainly to regions involved in self-referential activity and reward processing, with robust anatomical overlap with areas of the DMN. The more lenient approach of evaluating the overlap with canonical networks from an independent rs-fMRI dataset (Shirer et al. 2012) confirmed that this extended network mapped onto the DMN but also, although modestly, onto the salience network. Component 2, explaining only 8.7% of variance in the data instead appears to map mainly to regions involved in sensorimotor processing. The results for the FuncSurg targets showed that most of the variance present in the underlying functional networks was explained by 4 components; in this case, the variance was more dispersed (from 12 to 37%). Here, the large-scale meta-analytic decoding with Neurosynth showed that all 4 rs-fMRI-derived PCA networks mapped robustly onto areas involved in sensorimotor cognition (components 1,2,3, and 4) and onto the sensorimotor areas (components 1,3, and 4), with a preferential anatomical overlap with regions of the motor system, albeit to varying degrees. The more lenient approach evaluating the overlap with the canonical networks from an independent rs-fMRI dataset (Shirer et al. 2012), supported this conclusion but also indicated that all these networks significantly overlap with the salience network.

Based on the previous premise, overall, these results indicate that the targets of rt-fMRI-NF studies informed by the funcSurg approaches should be sought mainly among the main hubs of the sensorimotor network, possibly in the motor system and, although less robustly supported, among regions of the salience network, such as the aIns and the MCC already targeted by the previous rt-fMRI-NF studies. It is important to emphasize that the motor cortex is a key target of invasive (motor cortex stimulation) and non-invasive (transcranial magnetic stimulation or direct transcranial stimulation) approaches to controlling chronic pain. Although further studies are needed to assess its efficacy, a recent clinical study has shown that the (invasive) stimulation of the motor cortex can reduce the level of pain in a considerable number of neuropathic pain patients (Hamani et al. 2021). Similarly, noninvasive stimulation methods on primary motor areas can induce analgesic effects, perhaps through the restoration of defective endogenous pain inhibitory pathways (Giannoni-Luza et al. 2020); however, their effects appear to be short-lived (Nguyen et al. 2011). In support of the idea that the motor cortex is an important target for the control of chronic pain, a recent seminal study (Gan et al. 2022) showed that activation of neurons in layer 6 of M1 reduces the negative emotional valence and the related behavioral correlates of neuropathic pain, due to a connection between M1 and the nucleus accumbens (through a mid-dorsal thalamus pathway).

Although less supported by our results, since it emerges only with the canonical rs-fMRI networks approach, the salience network can also be considered a target of the rt-fMRI-NF. The salience network, anchored in the aIns and the MCC (Seeley et al. 2007) plays an important role in the guidance of flexible behavior and detecting salient events in the environment (Menon and Uddin 2010; Seeley et al. 2007; Yao et al. 2018b) integrating the current interoceptive state and autonomic feedback with internal goals and external demands (Craig 2009; Yao et al. 2018a). Recently, the salience network was shown to robustly overlap with the central autonomic network (Ferraro et al. 2022a). Moreover, abnormalities of the salience network have been repeatedly observed in chronic pain, with a recent meta-analysis (Ferraro et al. 2022b) showing robust dysregulations of the left aIns in this condition. Interestingly, the uniform map obtained by Neurosynth for the term “chronic pain” (along with the term “pain”) overlaps robustly with regions of the salience network, but not with regions of the sensorimotor network. This intriguing dissociation between the neural processing of chronic pain and the antinociceptive effects induced by the modulation of funcSurg targets (mapping onto the sensorimotor network and less robustly to the salience network) suggests that the neural pathways of these linked processes might be fundamentally different.

Notably, closely related to the observation that all functional networks underlying funcSurg targets map onto the sensorimotor and salience network, here we have also provided robust evidence that, despite a different involvement of additional neurotransmitter systems, 3 out of 4 networks underlying funcSurg targets map onto the noradrenergic system. This observation suggests that funcSurg approaches here investigated can regulate the noradrenergic system. This circuit is known to be involved in several cognitive functions (such as attention and vigilance), but also in the regulation of mood, anxiety, stress, and pain (Pertovaara 2013). Despite a long tradition of studies, the role of the noradrenergic system in the control of chronic pain has yet to be clarified, as both inhibitory and facilitatory effects have been observed. Recently, it has been proposed that with the passage of time after nerve injury, adrenergic neurons would switch from antinociceptive to pronociceptive, participating in the development of allodynia and hyperalgesia in neuropathic pain (Taylor and Westlund 2017). However, this contrasts with the fact that antidepressants capable of increasing monoamine levels are one of the main therapies used to treat chronic pain (Kuner and Kuner 2020).

### 4.1. Limitations

Despite interesting results, this work is clearly limited. First of all, the extrapolation of coordinates from funcSurg studies, which did not report the exact coordinates in the MNI spaces, was a major challenge. To overcome this issue, three senior experts in neuroimaging data independently extracted the coordinates, and subsequently, after the verification that no identified coordinate varied by more than 5 mm by the first ones, the center of mass was computed, and it was cross-referenced with the in-vivo high-resolution structural MRI human thalamic atlas (Saranathan et al. 2021). Second, we relied heavily on rs-fMRI data, inherently characterized by low spatial resolution, to identify the underlying rs-fMRI networks. However, in an attempt to mitigate this limitation, we utilized the high-resolution neuroimaging HCP dataset obtained at 7T MRI (spatial resolution of 1.6mm isotropic voxels) and the CONN toolbox. Indeed, in the context of the employed analyses (seed-based), CONN utilizes smoothed data at each voxel to define each voxel time series while unsmoothed data across all voxels within each seed to define each seed time series. Consequently, we could have utilized 3mm radius ROIs when extracting the rs-fMRI networks of thalamic and PAG/PVG targets, despite applying a 6mm smoothing. In addition, it’s worth noting that the spatial extent of activation in thalamic DBS, with a stimulation amplitude ranging from 1 to 3.5 V, typically results in an average effective activation radius of 2.0 to 3.9 mm. This falls within the range of our chosen ROI size of 3 mm, underscoring the compatibility of the DBS activation radius with our ROI dimensions (Kuncel, Cooper, and Grill 2008). Furthermore, the clear emergence of 4 distinct PCA rs-fMRI-derived maps for the funcSurg targets supports our findings, indicating differences among the different targets. The fourth limitation is associated with our use of rs-fMRI data from healthy participants to understand the potential networks modulated in Func-Neuro studies investigating chronic pain patients. However, it’s noteworthy that while aberrant networks have been identified in various neurological and neuropsychiatric disorders, a seminal study has demonstrated that DBS of the subthalamic nuclei in Parkinson’s disease can restore the activity of the stimulated network (motor network) to levels similar to those observed in healthy participants (Adam et al. 2022). As a note of caution, because the funcSurg studies considered here mainly investigated patients with neuropathic pain, we do not know the extent to which these results apply to patients suffering from other types of chronic pain.

### 4.2. Conclusion

Informed by the long tradition of funcSurg studies, our findings robustly support that the rt-fMRI-NF targets should be identified in areas of the motor system and, although less robustly, into areas of the salience network. Since areas of the salience network were already investigated with inconsistent findings, future works should investigate with rigorous clinical trials whether the motor system is a suitable target for the rt-fMRI-NF for the treatment of chronic pain, in particular in neuropathic pain patients.

Importantly, our results also suggest that some of the antinociceptive effects of the funcSurg approaches could be linked to the restoration of possible abnormal activity of the noradrenergic system.

## Data Availability

All data produced in the present study are available upon reasonable request to the authors

## Funding sources

This work was supported by Key R&D project of Science and Technology Department of the Sichuan Province (China), Grant number M112022YFWZ0003.

## Conflict of interest

The Authors report no conflict of interest.

## Acknowledgments

The authors would like to thank Prof. Ludovico Minati for his contribution to the identification of target coordinates extracted from the functional neurosurgery studies and Dr. Vincent Bazinet for his insights into methodological aspects.

## Disclaimer

Any opinions, findings, conclusions, or recommendations expressed in this publication do not reflect the views of the Government of the Hong Kong Special Administrative Region or the Innovation and Technology Commission.

## Notes

### Competing Interest Statement

The authors have declared no competing interest.

### Author Declarations

Young Adult HCP dataset (Smith et al., 2013; Van Essen et al., 2013), for details, see https://www.humanconnectome.org/hcp-protocols-ya-7t-imaging); http://findlab.stanford.edu/research.html (Shirer et al., 2012)

